# Identification and impact of microbiota-derived metabolites in ascites of ovarian and gastrointestinal cancer

**DOI:** 10.1101/2025.04.03.25325183

**Authors:** Sisi Deng, Wooyong Kim, Kefan Cheng, Qianlu Yang, Yogesh Singh, Gyuntae Bae, Nicolas Bézière, Lukas Mager, Stefan Kommoss, Jannik Sprengel, Christoph Trautwein

## Abstract

**Background:** Malignant ascites is a common complication of advanced ovarian and gastrointestinal cancer, significantly affecting metastasis, patient quality of life, and survival. In here, increased intestinal permeability cannot only result in blood or lymph infiltration but also microbial translocation from the gastrointestinal or uterine tract. This study aimed to discover microbiota-derived metabolites in ascites of ovarian cancer stages II-III, IV, and gastrointestinal cancer and to assess and discuss their potential roles in tumor progression and aggressiveness.

**Methods:** In an explorative approach, this study analyzed malignant ascites samples from a total of 18 ovarian and gastrointestinal cancer patients. Using reversed phase (RP) and hydrophilic interaction liquid chromatography (HILIC) coupled to trapped ion mobility time of flight mass spectrometry (timsTOF), we performed a fully untargeted 4D metabolomics approach. Additionally, a targeted flow cytometry-based cytokine panel was used to screen for inflammatory markers. Uni-and multivariate statistics were applied to identify significantly altered metabolites and lipids between cancer types and tumor stages. Non-endogenous, and thereof potentially microbiota-derived metabolites were identified using the Human Microbial Metabolome Database (MiMeDB).

**Results:** Distinct metabolic differences were observed between ovarian cancer (OC) stages II-III and gastrointestinal cancer (GI) groups, while stage IV OC showed metabolic profiles similar to GI cancers. In GI patients, microbiota derived metabolites showed higher levels of 3-methylindole, 3-methylxanthine, caffeine, D-glucurono-6,3-lactone, D-tagatose, glucosamine, LPCs, and trimethylamine N-oxide, with lower levels of benzamide, phosphocholine, sphinganine, and thymol compared to OC. Stage IV patients exhibited elevated concentrations of 1-methylhistidine, 3-hydroxyanthranilic acid, 4-pyridoxic acid, biliverdin, butyryl-L-carnitine, indole, LPI 18:1, mevalonic acid, and phenol, with reduced levels of naringenin, o-cresol, and octadecanedioic acid compared to OC stage II-III. Correlation analysis with cytokines showed a positive correlation of glucosamine, D-tagatose, trimethylamine N-oxide, caffeine, LPC 22:4, and LPC 20:1 with IL-10, while benzamide and thymol negatively correlated. Furthermore benzyl alcohol, naringenin, and phenol correlated positively with MCP-1, while 1-methylhistidine, 4-pyridoxic acid, and mevalonic acid showed negative correlations. These altered metabolites can be mostly linked to lipid metabolism and inflammatory pathways.

**Conclusion:** Dysregulated lipid metabolism plays a crucial role in ovarian cancer progression. Furthermore, 3-hydroxyanthranilic acid, indole, and naringenin indicate heightened inflammation and metabolic stress, serving as potential markers of disease progression. Deep metabolic phenotyping of ascites by timsTOF based 4D metabolomics elucidated the presence of microbiota-derived metabolites in ascites and distinct metabolic signatures in OC and GI. Involvement in typical tumor-related pathways pinpoints the relevance of these metabolites in the interaction between gut microbiota, the tumor microenvironment, and cancer biology and underscores the significant influence of microbiota in shaping malignant ascites.

## Introduction

Ovarian cancer (OC) is a leading cause of cancer-related mortality among women and is often termed the "silent killer" due to its asymptomatic nature in the early stages and lack of effective screening tools(1, 2). Consequently, most OC cases are diagnosed at advanced stages when the disease has metastasized, limiting treatment options and resulting in a poor prognosis(3). A common feature of advanced OC is the abnormal accumulation of fluid in the abdominal cavity, known as ascites, which significantly contributes to patient mortality(4). Ascites not only serves as a clinical hallmark of OC but is also observed in other malignancies such as gastrointestinal (GI) cancers and cirrhosis, and is rarely associated with non-cancerous conditions like heart failure and peritoneal tuberculosis(5, 6).

The pathogenesis of ascites is multifactorial, involving hypoalbuminemia, increased vascular permeability, impaired lymphatic drainage, and fluid retention due to activation of the renin–angiotensin–aldosterone system (RAAS)(7-9). Recent research has highlighted the role of the gut microbiota in modulating these processes through its influence on immune and metabolic pathways. For instance, butyrate produced by gut bacteria can inhibit RAAS activity, reducing fluid retention(10-12). Disruption of gut microbial balance in advanced disease may exacerbate intestinal permeability and promote bacterial translocation(13-17), contributing to ascites formation and creating a vicious cycle of inflammation and fluid accumulation.

One of the main complications of ascites is bacterial peritonitis (BP), which has been well-documented in patients with GI malignancies(18, 19). BP is often spontaneous and linked to bacterial translocation from the GI tract to the mesenteric lymph nodes(20). While less frequently reported in OC, the extensive metastasis and abdominal involvement in advanced OC may similarly compromise gastrointestinal integrity, increasing susceptibility to BP. The partially overlapping features of ascites in OC and GI cancers raise questions about the potential role of the microbiome in shaping the tumor-promoting properties of this fluid.

Moreover, ascites is not just a passive by-product of malignancy but may actively shape the tumor microenvironment, promoting metastasis and therapeutic resistance(21, 22). Its heterogeneous nature — ranging from clear and free-flowing to viscous and loculated — indicates distinct underlying biological processes that could influence tumor behavior. While paracentesis provides temporary symptomatic relief, it is often palliative and requires repeated procedures, carrying risks such as infection and protein loss. Despite its clinical significance, the molecular and microbial landscape of OC-associated ascites remains poorly understood.

In this study, we utilized LC-MS-based metabolomics and flow cytometry to characterize the metabolic and cytokine profiles of ascitic fluid, while also evaluating the potential impact of microbiota-derived metabolites. By dissecting these complex interactions, we aim to enhance our understanding of the pathophysiology of ascites and its contribution to OC and GI progression.

## Material and Methods

This exploratory study included 10 malignant ascites specimens from patients undergoing ovarian cancer (OC) resection and 8 malignant ascites specimens from Gastrointestinal (GI) cancers patients. Metabolomics and cytokine analysis were used to combine the results for in-depth phenotyping of malignant ascites.

### 1. Ethical Background

This study was conducted in accordance with the principles outlined in the Declaration of Helsinki and approved by the Ethics Committee, Faculty of Medicine, University of Tübingen, Germany (Ref. Nr. 696/2016BO2 and 117/2020BO1). Written informed consent was obtained from all participating patients. The collection of samples did not interfere with or alter patient treatment plans. All data were anonymized in compliance with the European General Data Protection Regulation (GDPR) and applicable German data protection laws.

### 2. Collection and Storage of Ascitic Fluid Samples

Ascites samples were collected from patients undergoing surgery for ovarian cancer at the Department of General and Transplant Surgery and the Women’s Hospital, University Hospital Tübingen, Germany. Samples were obtained under sterile conditions in the operating room, stored in sterile tubes, and immediately transported to the laboratory using an icebox to maintain low temperatures. Upon arrival, the samples were centrifuged at 4°C for 30 minutes at 1,200 rpm. The resulting supernatant was aliquoted into 2 mL tubes and stored at −80°C until further analysis. Relevant patient information, including demographic data, cancer histology, and the extent of peritoneal disease, was recorded. All samples and associated patient data were anonymized prior to analysis.

### 3. Quantification of Cytokines in Malignant Ascites

To quantify cytokine levels in malignant ascites, 25 µL of ascites from ovarian cancer patients was mixed with 25 µL of assay buffer. Next, 25 µL of a 13-plex bead mix from the LEGENDplex™ Human Inflammation Panel 1 (13-plex, #740809, BioLegend, USA) was added to each well of a 96-well microplate. This multiplex bead-based assay is capable of quantifying 13 different cytokines/chemokines with the following minimum detectable concentrations (MDC) in parentheses: IL-1β (1.5 ± 0.6 pg/mL), IFN-α2 (2.1 ± 0.2 pg/mL), IFN-γ (1.3 ± 1.0 pg/mL), TNF-α (0.9 ± 0.8 pg/mL), MCP-1 (1.1 ± 1.2 pg/mL), IL-6 (1.5 ± 0.7 pg/mL), IL-8 (2.0 ± 0.5 pg/mL), IL-10 (2.0 ± 0.5 pg/mL), IL-12p70 (2.0 ± 0.2 pg/mL), IL-17A (0.5 ± 0.0 pg/mL), IL-18 (2.0 ± 0.5 pg/mL), IL-23 (1.8 ± 0.1 pg/mL), and IL-33 (4.4 ± 1.5 pg/mL).

The plate was incubated and shaken at room temperature for 2 hours, allowing the analytes (cytokines) to bind to the corresponding antibody-conjugated capture beads. Following incubation, the wells were washed to remove unbound analytes. Biotinylated detection antibodies (25 µL) were then added and allowed to bind to the analyte-bound beads. After 30 minutes of incubation, 25 µL of streptavidin–phycoerythrin (SA-PE) was added, which binds to the biotinylated detection antibodies and produces a fluorescent signal proportional to the amount of each cytokine.

After a further 1-hour incubation, the beads were washed, resuspended in wash buffer, and analyzed using a flow cytometer. Fluorescent signals were measured, and the concentrations of the analytes were determined based on standard curves generated using the LEGENDplex™ data analysis software (BioLegend, USA).

### 4. Sample Preparation for timsTOF LC-MS Analysis

Frozen ascites samples were thawed at room temperature, and 1.2 mL of each sample was transferred into separate 1.5 mL Eppendorf tubes. The tubes were centrifuged at 14,000 rpm at 4°C for 15 minutes. The supernatants (1 mL) were carefully collected and evaporated at room temperature using a Concentrator Plus (Eppendorf, Wesseling-Berzdorf, Germany) for 6 hours.

#### 4.1. Reversed-Phase Liquid Chromatography (RPLC) Sample Preparation

For reversed-phase (RP) liquid chromatography analysis, the dried samples were reconstituted in 100 µL of MilliQ water (MQ) and 300 µL of ice-cold (-20°C) high-performance liquid chromatography (HPLC) grade acetonitrile (VWR Chemicals, Darmstadt, Germany). The mixture was vortexed for 1 minute, followed by incubation at -20°C for 10 minutes. Samples were then centrifuged at 14,000 rpm at 4°C for 15 minutes. From each tube, 300 µL of the supernatant was transferred to a new Eppendorf tube and evaporated to dryness for 1.5 hours at room temperature. The dried samples were then reconstituted in 60 µL of MQ/acetonitrile (9:1, v/v), vortexed for 10 seconds, and centrifuged again at 14,000 rpm at 4°C for 15 minutes. Finally, 50 µL of the supernatant was transferred to HPLC vials (VWR, Leuven, Belgium) equipped with inserts for subsequent RPLC-MS analysis.

#### 4.2. Hydrophilic Interaction Liquid Chromatography (HILIC) Sample Preparation

For hydrophilic interaction liquid chromatography (HILIC) analysis, the dried samples were reconstituted in 100 µL of MQ and 300 µL of ice-cold (-20°C) acetonitrile. The mixture was vortexed for 1 minute, followed by incubation at -20°C for 10 minutes. The samples were centrifuged at 14,000 rpm at 4°C for 15 minutes, and the resulting supernatant (300 µL) was transferred into HPLC vials with inserts for HILIC-MS measurements.

### 5. Liquid Chromatography Conditions

Analyte separation was performed using the Elute PLUS LC series (Bruker, Bremen, Germany).

#### 5.1. RPLC Conditions

RPLC separations were conducted on an Intensity Solo 2 C18 Column (100 Å; 2.0 µm; 2.1 mm × 100 mm; #BRHSC18022100, Bruker) using 0.1% formic acid in MilliQ water as mobile phase A and 0.1% formic acid in acetonitrile as mobile phase B. A 5 µL injection of each sample was used. The separation was carried out at a flow rate of 0.4 mL/min from 0 to 9 min and 10.6 to 13 min, and at 0.6 mL/min from 9.1 to 10.6 min, with a column temperature maintained at 50°C using the following gradient: 0-1 min, 5% B; 1-7 min, 5-40% B; 7-9 min, 40-98% B; 9-10.6 min, 98% B; 10.6-10.7 min, 98-5% B; 10.7-13 min, 5% B.

#### 5.2. HILIC Conditions

HILIC separations were performed on an ACQUITY UPLC BEH Amide column (130 Å, 1.7 µm, 2.1 mm × 150 mm; #186004802, Waters) using 10 mM ammonium formate and 0.1% formic acid in MilliQ water as mobile phase A and 10 mM ammonium formate and 0.1% formic acid in acetonitrile as mobile phase B. A 5 µL injection was used, with separation conducted at a flow rate of 0.5 mL/min and a column temperature of 40°C using the following gradient: 0-3 min, 100% B; 3-10 min, 100-85% B; 10-14 min, 85-50% B; 14-15 min, 50% B; 15-15.1 min, 50-100% B; 15.1-23 min, 100% B.

### 6. Mass Spectrometry Analysis

The separated analytes were analyzed using a timsTOF fleX mass spectrometer (Bruker, Bremen, Germany) equipped with an Apollo II source for RP measurements, and a timsTOF Pro 2 (Bruker, Bremen, Germany) with a vacuum-insulated probe heated electrospray ionization (VIP-HESI) source for HILIC analysis. LC-MS/MS data were acquired in duplicate (technical replicates) using positive and negative PASEF modes, with a TOF mass range of m/z 20-1300. The system was controlled using timsControl® and Compass HyStar® software, and data acquisition was managed using the same software. Quality control (QC) samples were run every ten injections, and blank samples were analyzed using H₂O for RP and acetonitrile for HILIC.

### 7. Data Preprocessing and Statistical Analysis

Raw data processing was conducted using MetaboScape® software (version 2024b, Bruker) with four-dimensional feature extraction, capturing mass-to-charge ratio (m/z), isotopic pattern quality, retention times, MS/MS spectra, and collision cross-section (CCS) values. Feature extraction was performed using the T-ReX® 4D algorithm, followed by annotation through the Bruker Human Metabolome Database (HMDB) and the NIST Mass Spectral Library. High-quality spectra were selected based on stringent criteria, including chromatogram and ion mobilogram quality, annotation scores, and CCS accuracy.

Data from RP) and HILIC measurements were combined, and the final dataset was analyzed and visualized using MetaboAnalystR, Pheatmap, and ComplexHeatmap packages in R (version 4.3.2). Sample intensities were normalized using probabilistic quotient normalization (PQN) and log-transformed. Descriptive statistics and correlations were calculated in R, with comparative statistics conducted via t-tests and one-way analysis of variance (ANOVA) for normally distributed data, and non-parametric tests for skewed data.

## Results

### 1. Patients clinical Characteristics

Ascites samples were collected from 10 OC and 8 GI patients undergoing open surgery for malignancy removal. The clinicopathological characteristics of the patients are summarized in **Table 1**. None of the patients had received chemotherapeutic treatment before ascites collection. Notably, the OC patient with clear cell carcinoma did not have a history of endometriosis.

**Table 1.**
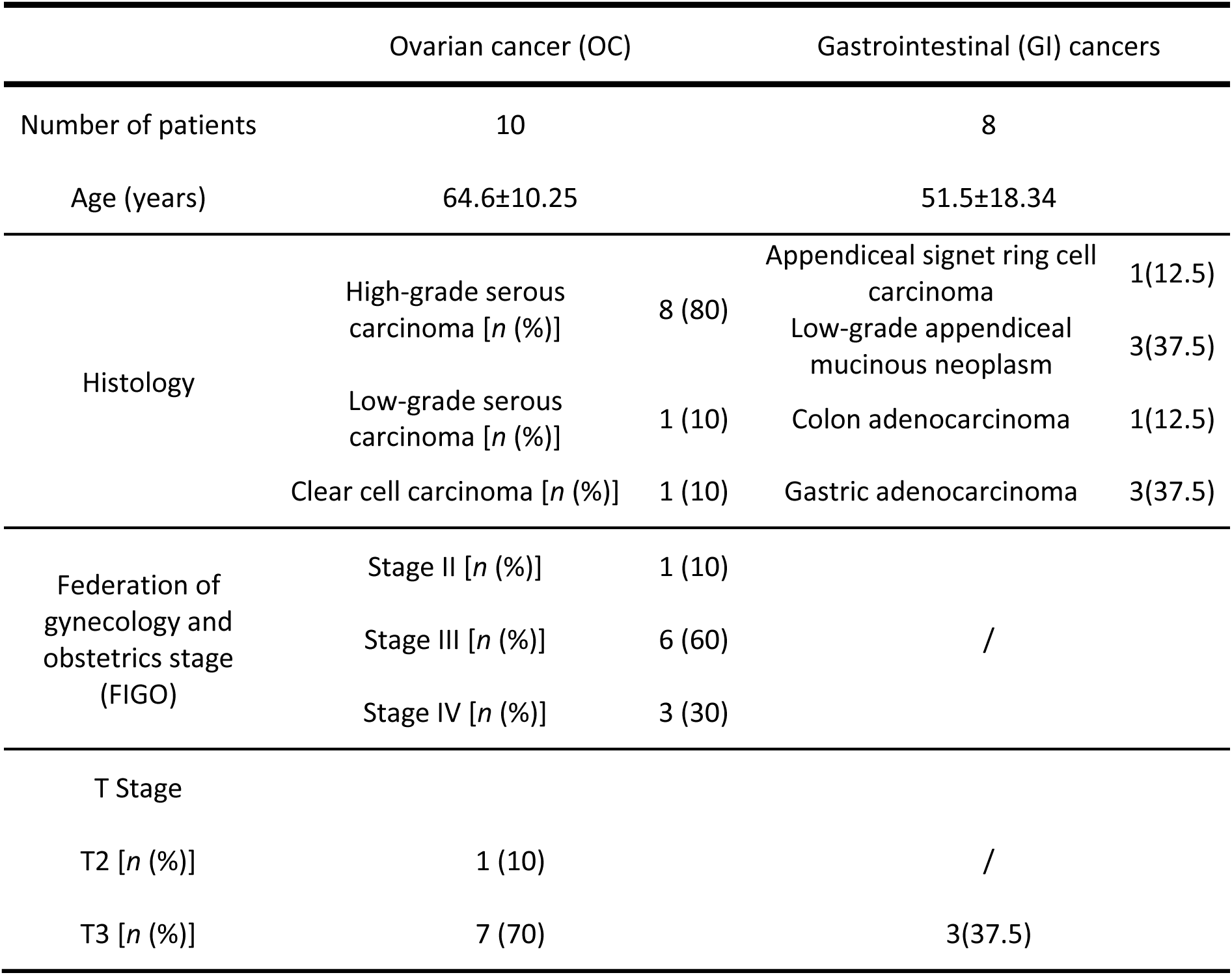
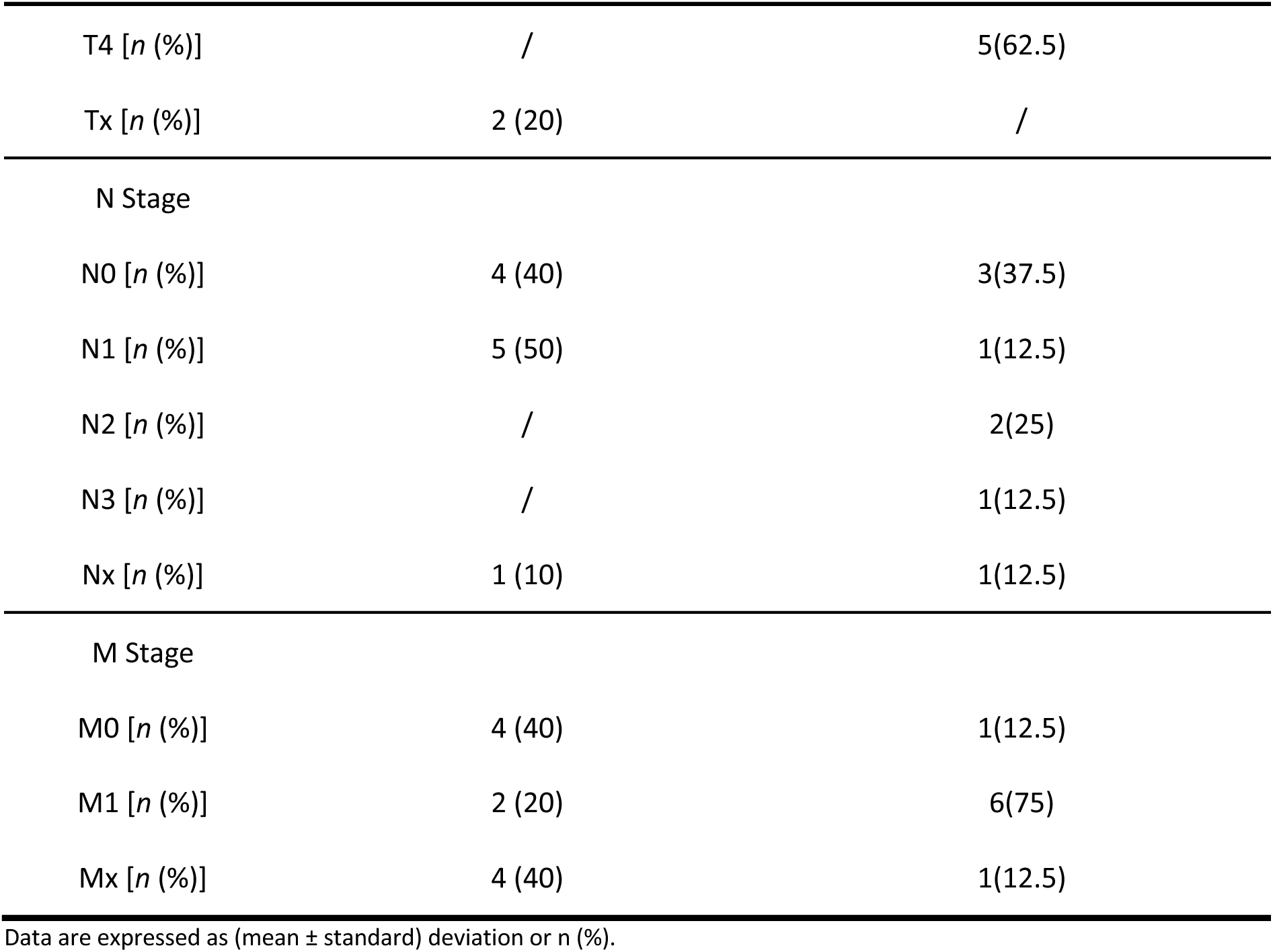
Clinic-pathological characteristics of the explorative cohort.

|  |  | Ovarian cancer (OC) | Gastrointestinal (GI) cancers |  |
| --- | --- | --- | --- | --- |
| Number of patients |  | 10 | 8 |  |
| Age (years) |  | 64.6±10.25 | 51.5±18.34 |  |
| Histology | High-grade serous carcinoma [*n* (%)] | 8 (80) | Appendiceal signet ring cell carcinoma | 1(12.5) |
|  |  |  | Low-grade appendiceal mucinous neoplasm | 3(37.5) |
|  | Low-grade serous carcinoma [*n* (%)] | 1 (10) | Colon adenocarcinoma | 1(12.5) |
|  | Clear cell carcinoma [*n* (%)] | 1 (10) | Gastric adenocarcinoma | 3(37.5) |
| Federation of gynecology and obstetrics stage (FIGO) | Stage II [*n* (%)] | 1 (10) | / |  |
|  | Stage III [*n* (%)] | 6 (60) |  |  |
|  | Stage IV [*n* (%)] | 3 (30) |  |  |
| T Stage |  |  |  |  |
| T2 [*n* (%)] |  | 1 (10) | / |  |
| T3 [*n* (%)] |  | 7 (70) | 3(37.5) |  |

|  |  |  |
| --- | --- | --- |
| T4 [n (%)] | / | 5(62.5) |
| Tx [n (%)] | 2 (20) | / |
| N Stage |  |  |
| N0 [n (%)] | 4 (40) | 3(37.5) |
| N1 [n (%)] | 5 (50) | 1(12.5) |
| N2 [n (%)] | / | 2(25) |
| N3 [n (%)] | / | 1(12.5) |
| Nx [n (%)] | 1 (10) | 1(12.5) |
| M Stage |  |  |
| M0 [n (%)] | 4 (40) | 1(12.5) |
| M1 [n (%)] | 2 (20) | 6(75) |
| Mx [n (%)] | 4 (40) | 1(12.5) |

Data are expressed as (mean ± standard) deviation or n (%).

### 2. Metabolite Differences Between Ovarian Cancer (Stages II-III, IV) and Gastrointestinal Cancer Groups

Distinct metabolite changes were observed between OC stages II-III and GI groups, while ovarian cancer stage IV samples displayed metabolic profiles similar to those of the GI cancer samples. Normalized intensity data from three groups—OC II-III (ovarian cancer stages II-III), OC IV (ovarian cancer stage IV), and GI (including appendiceal, colon, and gastric cancers)— were analyzed using one-way ANOVA with a false discovery rate (FDR) cutoff of 0.05. Twelve significant metabolites were identified out of the 696 measured. Approximately 92% of the metabolites showed distinct intensity distributions when comparing the GI group with the OC II-III group. However, when comparing the GI group with OC IV, around 83% of the metabolites exhibited similar intensity trends. A heatmap was generated to visualize the differences in normalized intensity levels for these significant metabolites across the three groups, as shown in **Figure 1**.

**Figure 1.**
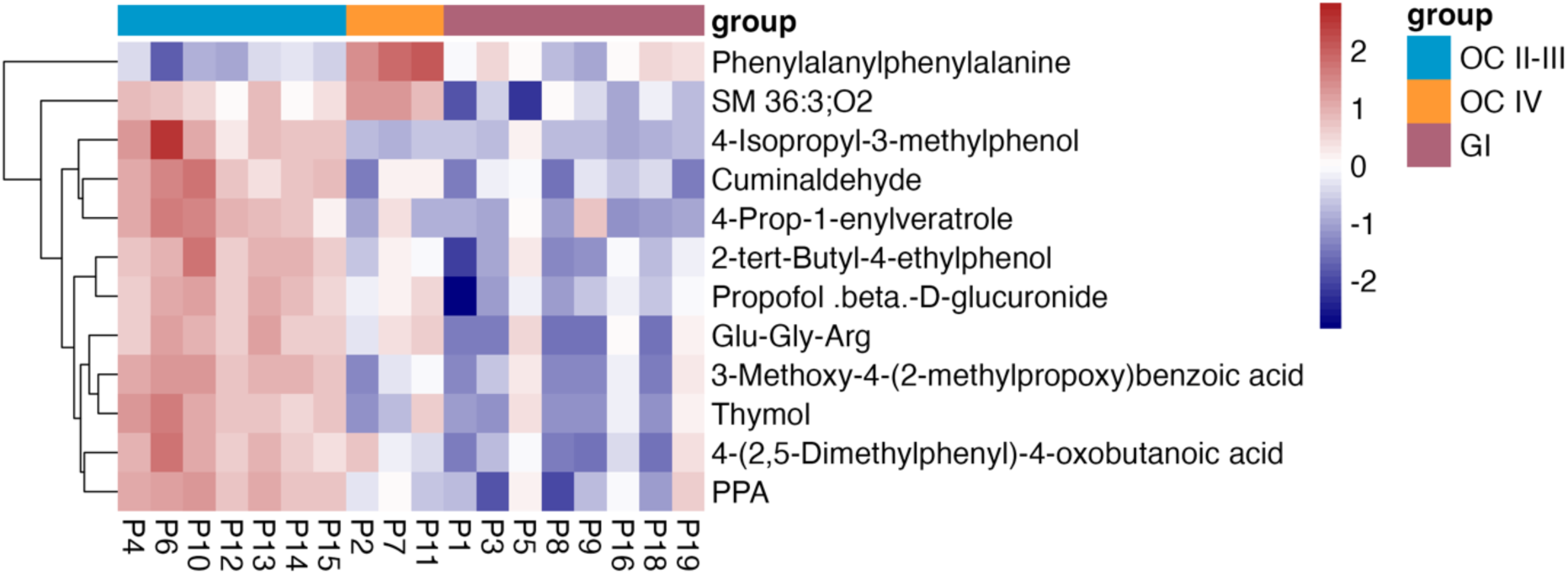
Identification of Ascites Metabolic Signatures in OC II-III, OC IV, and GI Groups. The heatmap shows 12 significant metabolites (ANOVA, adjusted p-value cutoff of 0.05) across eight GI, seven OC II-III, and three OC IV biological replicates. Clustering was performed using Ward’s hierarchical method, with Euclidean distance as the distance metric. Abbreviations: SM, sphingomyelin; Glu-Gly-Arg, glutamyl-glycyl-arginine; PPA, phenylpropionic acid.

The top 12 significant metabolites identified in the heatmap are further illustrated in individual raincloud plots, as shown in **Figure 2**.

**Figure 2.**
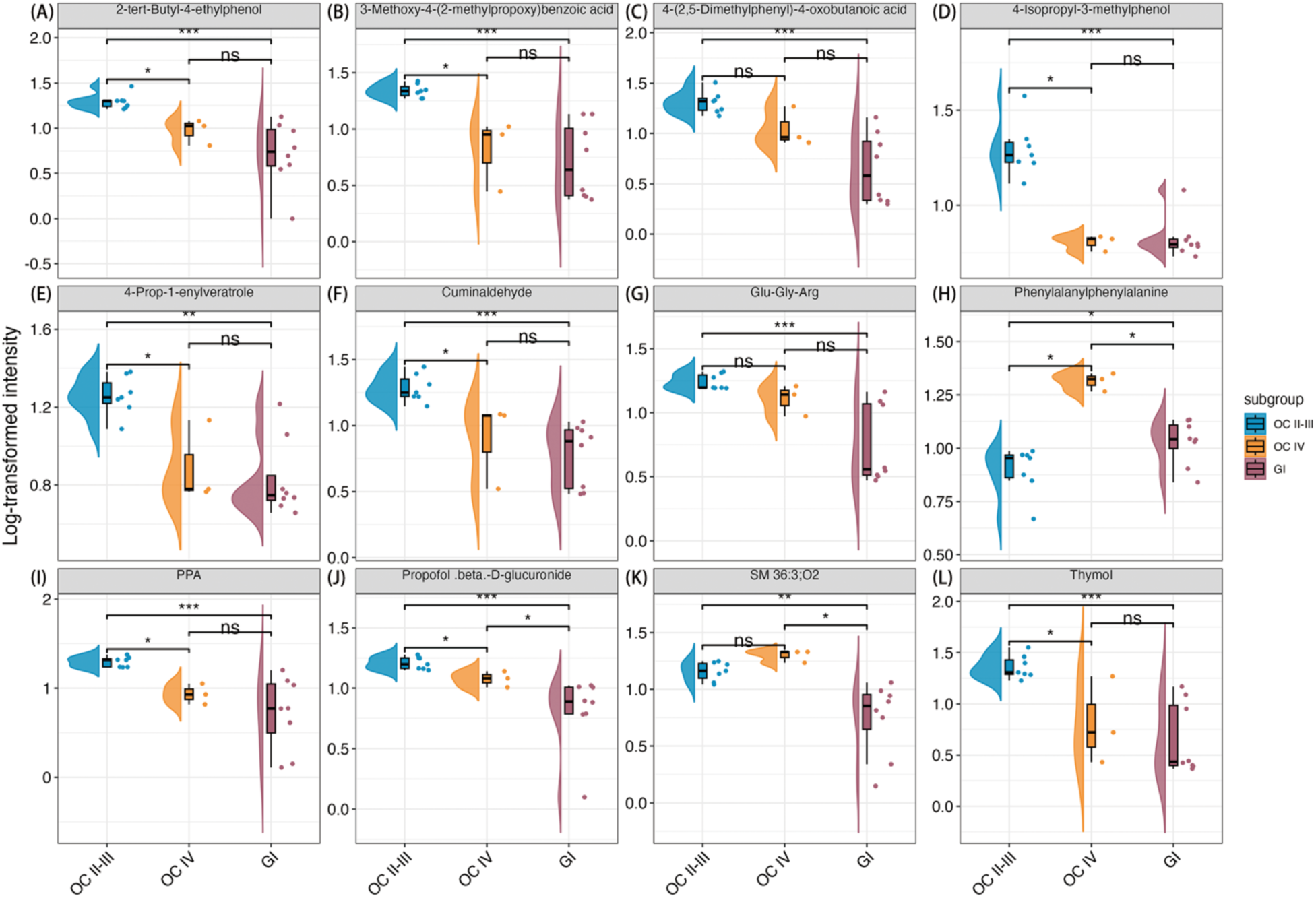
Raincloud Plots of Significantly Differential Metabolites. Raincloud plots (combined violin, box, and strip plots) display the significantly different metabolites: (A) 2-tert-butyl-4-ethylphenol, (B) 3-methoxy-4-(2-methylpropoxy)benzoic acid, (C) 4-(2,5-dimethylphenyl)-4-oxobutanoic acid, (D) 4-isopropyl-3-methylphenol, (E) 4-prop-1-enylveratrole, (F) cuminaldehyde, (G) Glu-Gly-Arg, (H) phenylalanylphenylalanine, (I) PPA, (J) Propofol-β-D-glucuronide, (K) SM 36:3;O2, and (L) thymol, comparing gastrointestinal cancer (GI), ovarian cancer stage II-III (OC II-III), and ovarian cancer stage IV (OC IV) groups. Pairwise t-tests were used for comparisons (*p < .05, **p < .01, ***p < .001, ****p < .0001). Dots represent individual data points, the middle line of the box represents the median, and the upper and lower ends of the box represent the upper and lower quartiles. Half-violins illustrate the data distributions. Abbreviations: SM, sphingomyelin; Glu-Gly-Arg, glutamyl-glycyl-arginine; PPA, phenylpropionic acid.

Ten metabolites exhibit similar metabolic trends. Among them, propofol-β-D-glucuronide (J) shows a gradual and significant decrease in a stepwise manner across the OC II-III, OC IV, and GI groups (OC IV significantly decreased compared to OC II-III, and GI significantly decreased compared to both OC II-III and OC IV).

Several metabolites, including 2-tert-butyl-4-ethylphenol (A), 3-methoxy-4-(2-methylpropoxy)benzoic acid (B), 4-isopropyl-3-methylphenol (D), 4-prop-1-enylveratrole (E), cuminaldehyde (F), PPA (I), and thymol (L), show similar trends. The OC IV group shows a significant decrease compared to OC II-III, and the GI group shows a significant decrease compared to OC II-III. While the GI group also decreases compared to OC IV, this difference is not significant. Similarly, 4-(2,5-dimethylphenyl)-4-oxobutanoic acid (C) and Glu-Gly-Arg (G) exhibit a decrease, but the reduction is only significant when comparing GI to OC II-III. The decreases between OC IV and OC II-III, and GI and OC IV, are not statistically significant.

Phenylalanylphenylalanine (H) and SM 36:3;O2 (K) follow a different trend. Phenylalanylphenylalanine (H) significantly increases in OC IV compared to both OC II-III and GI groups, and is also significantly elevated in the GI group compared to OC II-III. SM 36:3;O2 (K) follows the same pattern, though the difference between OC II-III and OC IV is not significant.

### 3. Potential Microbiota-derived Metabolites in Ascites Samples

#### 3.1. Identification and grading of non-endogenous metabolites with the Human Microbial Metabolome Database

The Human Microbial Metabolome Database (MiMeDB) (https://mimedb.org) is a comprehensive multi-omic resource for microbiome research(23). To explore the potential origins of metabolites that cannot be of human-origin and that were identified significant in OC II-III, OC IV, and GI ascites samples, a comparative analysis was first conducted between different stages of OC and between OC and GI. Using the MiMeDB database, significantly increased or decreased metabolites were then examined for their potential origin and relationship to microbiota species.

#### 3.2. Differential general and microbiome-derived metabolites in ascites of OC and GC

##### 3.2.1. Identification of general metabolite changes between OC and GI ascites

Based on a fold change (FC) > 1.2 and p-value < 0.05, the analysis of normalized intensity data between the OC and GI groups identified 90 significant metabolites out of the 696 measured. Of these, 51 metabolites showed significantly lower intensity, and 39 showed significantly higher intensity in the GI group compared to the OC group. Due to the small sample size of ascites samples, the raw p-value was used for this comparison instead of the FDR-adjusted p-value. The corresponding volcano plot is shown in **Figure 3**, where red dots represent metabolites with higher normalized intensity in the GI group, and blue dots represent those with lower intensity compared to the OC group.

**Figure 3.**
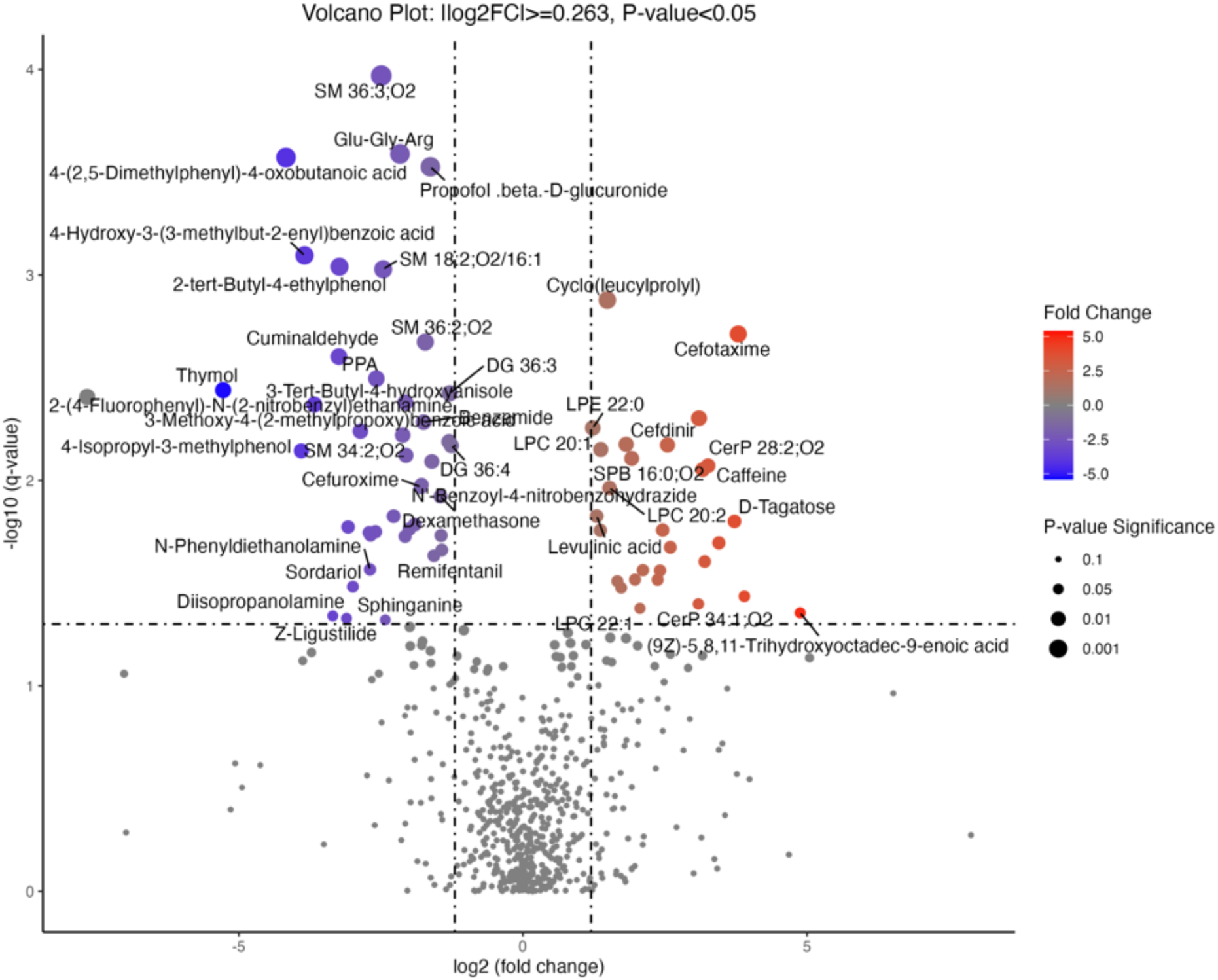
Volcano Plot Showing Statistically Significant Metabolite Changes Between OC and GI Groups. Red dots represent metabolites upregulated in the GI group, while blue dots represent those upregulated in the OC group. Fold change threshold: 1.2, raw p-value threshold: 0.05. Abbreviations: LPC, lysophosphatidylcholine; CerP, Ceramide phosphate; LPE, lysophosphatidylethanolamine; PE, Phosphatidylethanolamine; SPB, Sphingoid base; SM, sphingomyelin; PC, Phosphatidylcholine; DG, Diacylglycerol; Cer, Ceramide; DGTS, Diacylglyceryl-N,N,N-trimethylhomoserine; PPA, phenylpropionic acid; Glu-Gly-Arg, glutamyl-glycyl-arginine.

Sphinganine and phosphocholine were significantly decreased in the GI group compared to the OC group, while LPC 18:1, LPC 20:2, LPC 22:1, and LPC 22:4 were significantly increased in the OC group compared to the GI group. These changes suggest a disruption in lipid metabolism.

**Figure S1.**
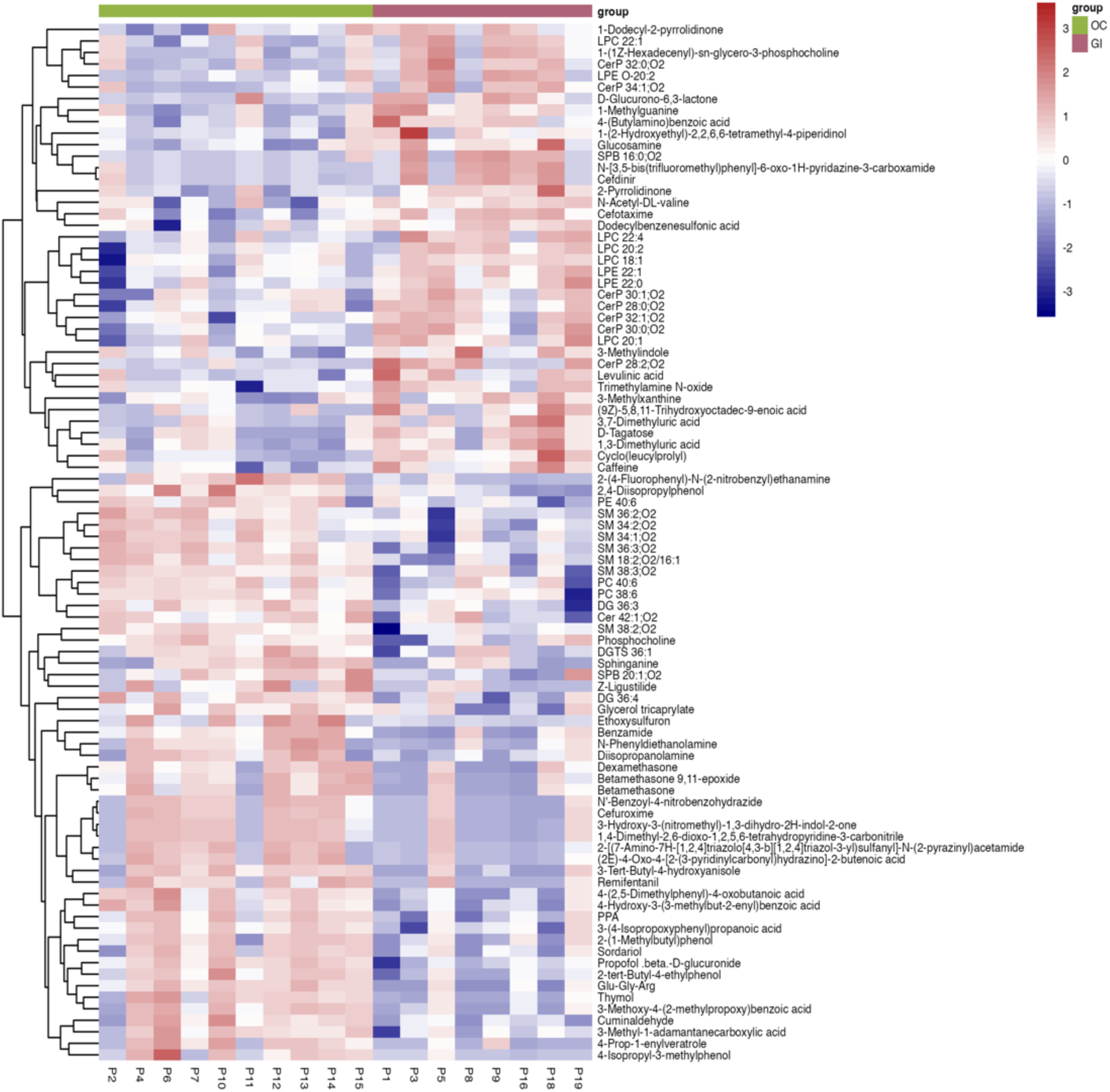
Identification of Ascites Metabolic Signatures in OC and GI Groups. The heatmap displays 90 significant metabolites (t-test, raw p-value cutoff of 0.05, fold change threshold of 1.2) across ten OC and eight GI biological replicates. Clustering was performed using Ward’s hierarchical method with Euclidean distance as the distance metric. Abbreviations: LPC, lysophosphatidylcholine; CerP, Ceramide phosphate; LPE, lysophosphatidylethanolamine; PE, Phosphatidylethanolamine; SPB, Sphingoid base; SM, sphingomyelin; PC, Phosphatidylcholine; DG, Diacylglycerol; Cer, Ceramide; DGTS, Diacylglyceryl-N,N,N-trimethylhomoserine; PPA, phenylpropionic acid; Glu-Gly-Arg, glutamyl-glycyl-arginine.

##### 3.2.2. Identification of microbiota-derived metabolite changes between OC and GI ascites

The MiMeDB database results identified a set of metabolites potentially produced or synthesized by the gut microbiome, including bacterial species, Eukaryota/Fungi, and Archaea. The associated phyla for the GI vs. OC comparison are listed in **Table 2**.

**Table 2.**
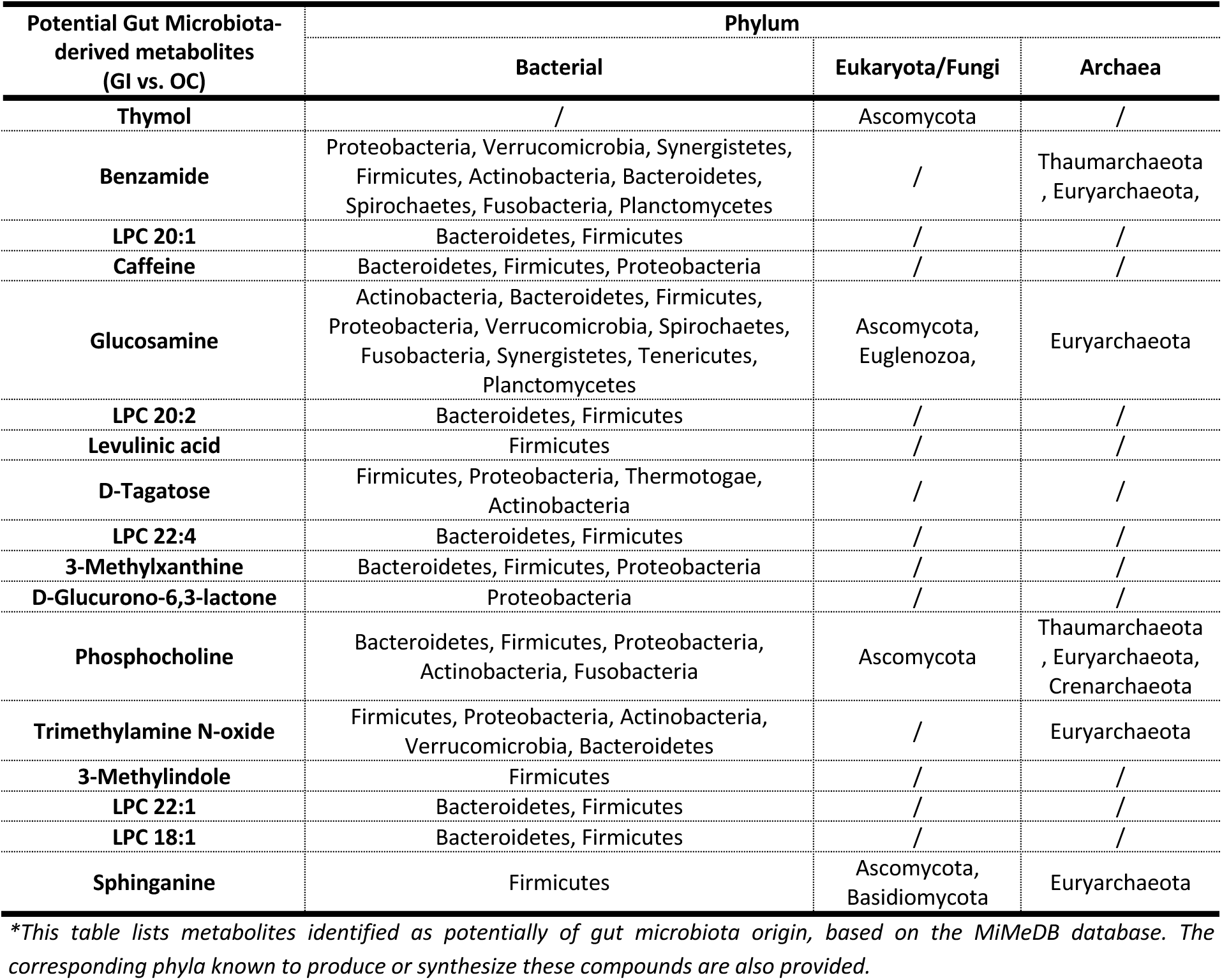
Metabolites Potentially Derived from the Gut Microbiota Identified in the Comparison Between Ovarian Cancer (OC) and Gastrointestinal Cancer (GI) Groups.

The differentiated potentially microbiota-derived metabolites are visualized in **Figure 4**. Metabolites that were significantly increased in GI ascites samples compared to the OC group include 3-methylindole (A), 3-methylxanthine (B), caffeine (D), D-glucurono-6,3-lactone (E), D-tagatose (F), glucosamine (G), levulinic acid (H), lysophosphatidylcholine 18:1 (LPC 18:1) (I), lysophosphatidylcholine 20:1 (LPC 20:1) (J), lysophosphatidylcholine 20:2 (LPC 20:2) (K), lysophosphatidylcholine 22:1 (LPC 22:1) (L), lysophosphatidylcholine 22:4 (LPC 22:4) (M), and trimethylamine N-oxide (Q). In contrast, benzamide (C), phosphocholine (N), sphinganine (O), and thymol (P) were significantly decreased in the GI group compared to the OC group.

**Figure 4.**
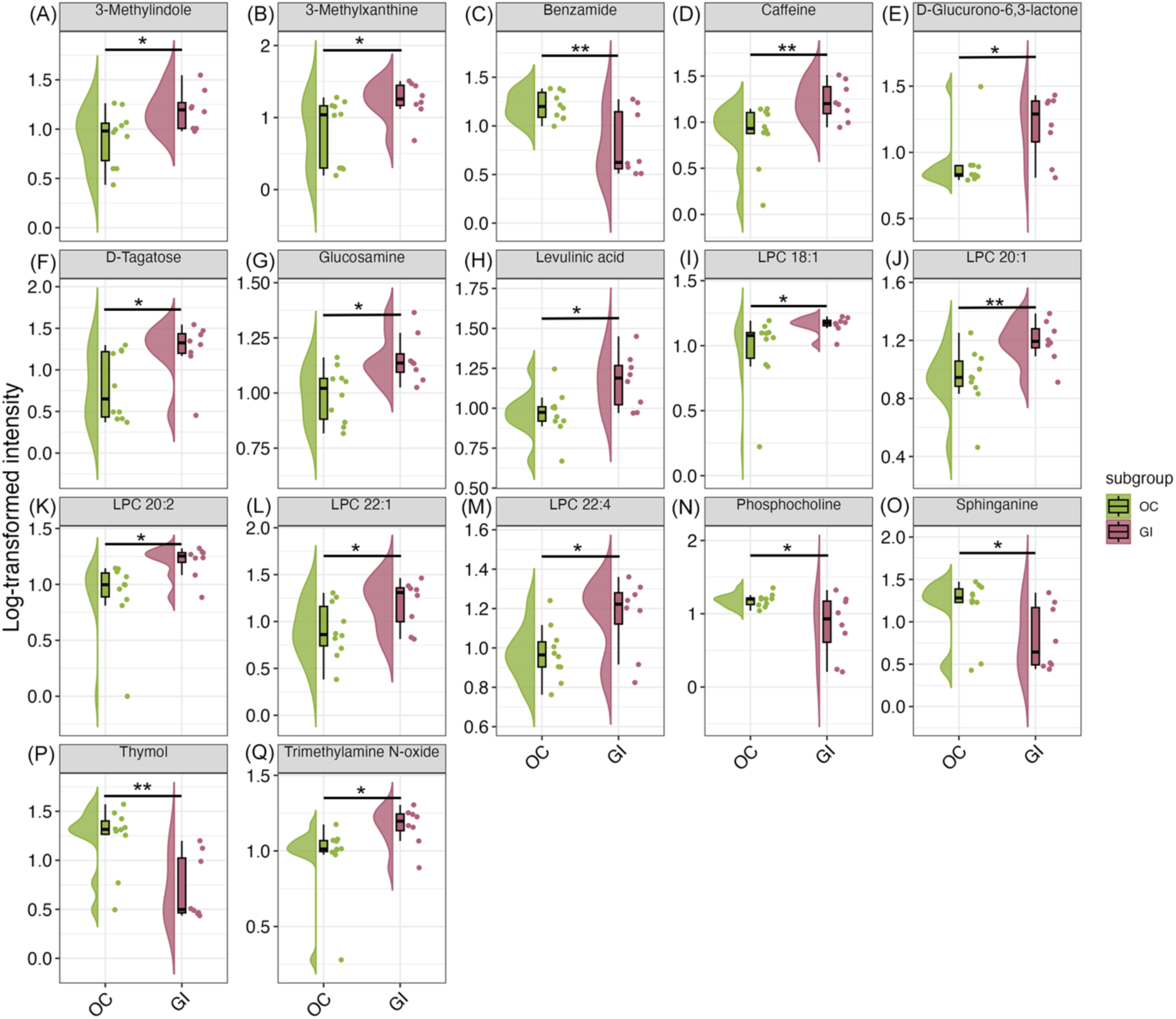
Raincloud Plots of Differentiated Potential Gut Microbiota-derived Metabolites Between OC and GI. Raincloud plots display the significantly different metabolites: (A) 3-methylindole, (B) 3-methylxanthine, (C) benzamide, (D) caffeine, (E) D-glucurono-6,3-lactone, (F) D-tagatose, (G) glucosamine, (H) levulinic acid, (I) lysophosphatidylcholine 18:1 (LPC 18:1), (J) lysophosphatidylcholine 20:1 (LPC 20:1), (K) lysophosphatidylcholine 20:2 (LPC 20:2), (L) lysophosphatidylcholine 22:1 (LPC 22:1), (M) lysophosphatidylcholine 22:4 (LPC 22:4), (N) phosphocholine, (O) sphinganine, (P) thymol, and (Q) trimethylamine N-oxide, comparing gastrointestinal cancer (GI) with ovarian cancer (OC) groups. Unpaired t-tests were used for comparisons with normalized parametric data (*p < .05, **p < .01, ***p < .001, ****p < .0001). Dots represent individual data points, the middle line of the box represents the median, and the upper and lower edges of the box represent the upper and lower quartiles. Half-violins illustrate data distributions. Abbreviations: LPC, lysophosphatidylcholine.

A circular heatmap (**Figure 5A**) was generated to visualize the overall changes in potential gut-derived metabolites across the 18 ascites samples from both the OC and GI groups. A corresponding Sankey plot (**Figure 5B**) illustrates the microbial sources and categories of the metabolites that increased or decreased in the GI and OC groups, including the associated phyla and their respective superkingdoms. Bacteria were the most predominant source, followed by Archaea, with Eukaryota/Fungi contributing the least.

**Figure 5.**
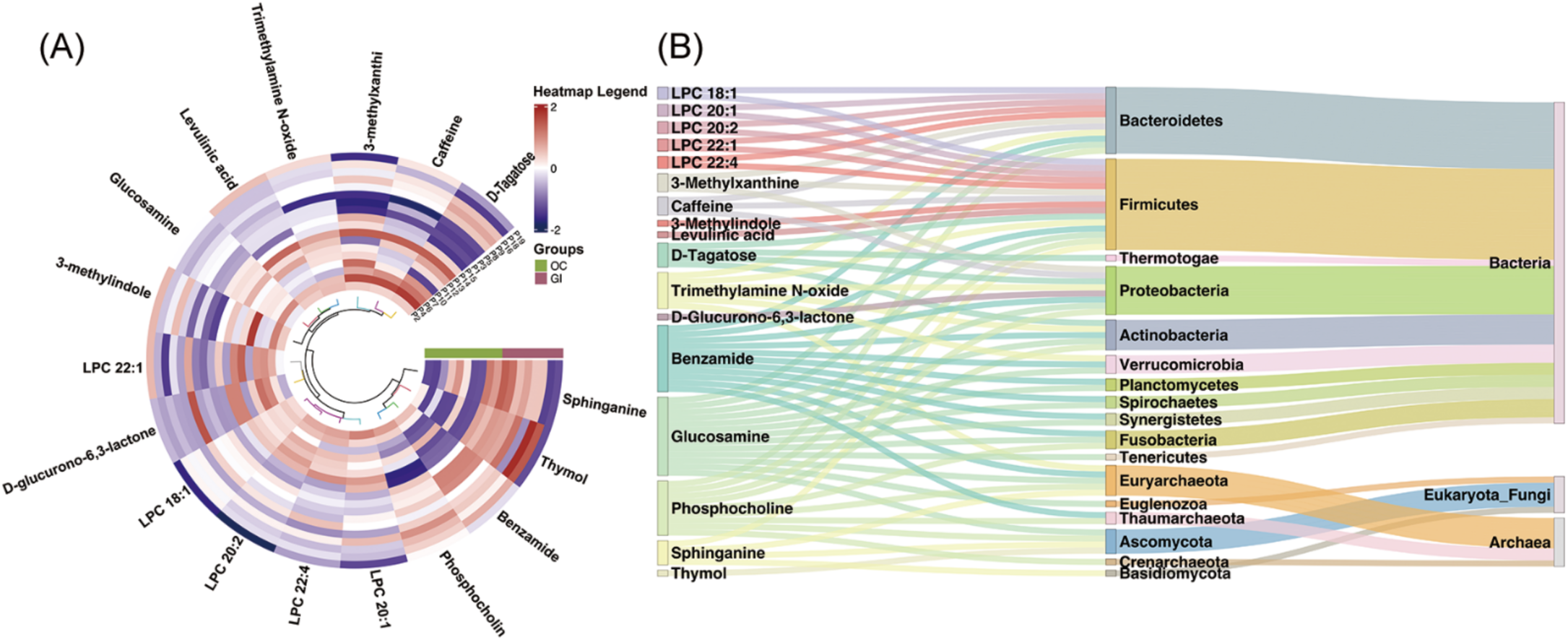
Intensity Differences of Potential Gut Microbiota-derived Metabolites Between GI and OC Groups and Their Microbial Sources and Classes. (A) The circular heatmap displays the intensity differences of 17 significant potential microbiome-derived metabolites across 10 OC and 8 GI biological replicates (t-test, raw p-value cutoff of 0.05, fold change threshold of 1.2). Clustering was performed using Ward’s hierarchical method with Euclidean distance as the distance metric. (B) The Sankey plot illustrates the structure of associated phyla and their respective superkingdoms for all differentiated potential gut microbiota-derived metabolites, highlighting the dominant role of gut bacteria in the ascites component of both ovarian cancer and gastrointestinal cancer. Abbreviations: LPC, lysophosphatidylcholine.

#### 3.3. Differential general and microbiome-derived metabolites in ascites of OC Stage II-III and Stage IV

#### 3.4. Identification of general metabolite changes between OC stage II-III and OC stage IV

A comparison between OC II-III (ovarian cancer stage II-III) and OC IV (ovarian cancer stage IV) identified 84 significant metabolites out of 649, with 45 showing significantly lower intensity and 39 showing significantly higher intensity in the OC IV group compared to OC II-III. This analysis was performed using a fold change threshold of 1.2 and a raw p-value threshold of 0.05. Due to the small sample size of the ascites samples, raw p-values were used instead of FDR-adjusted p-values. The corresponding volcano plot is shown in **Figure 6**, where red dots represent metabolites with higher normalized intensity in OC IV, and blue dots represent those with lower intensity compared to OC II-III. A heatmap of the 84 significant metabolites was generated to compare the metabolic profiles of OC II-III and OC IV, as shown in **Figure 6**.

**Figure 6.**
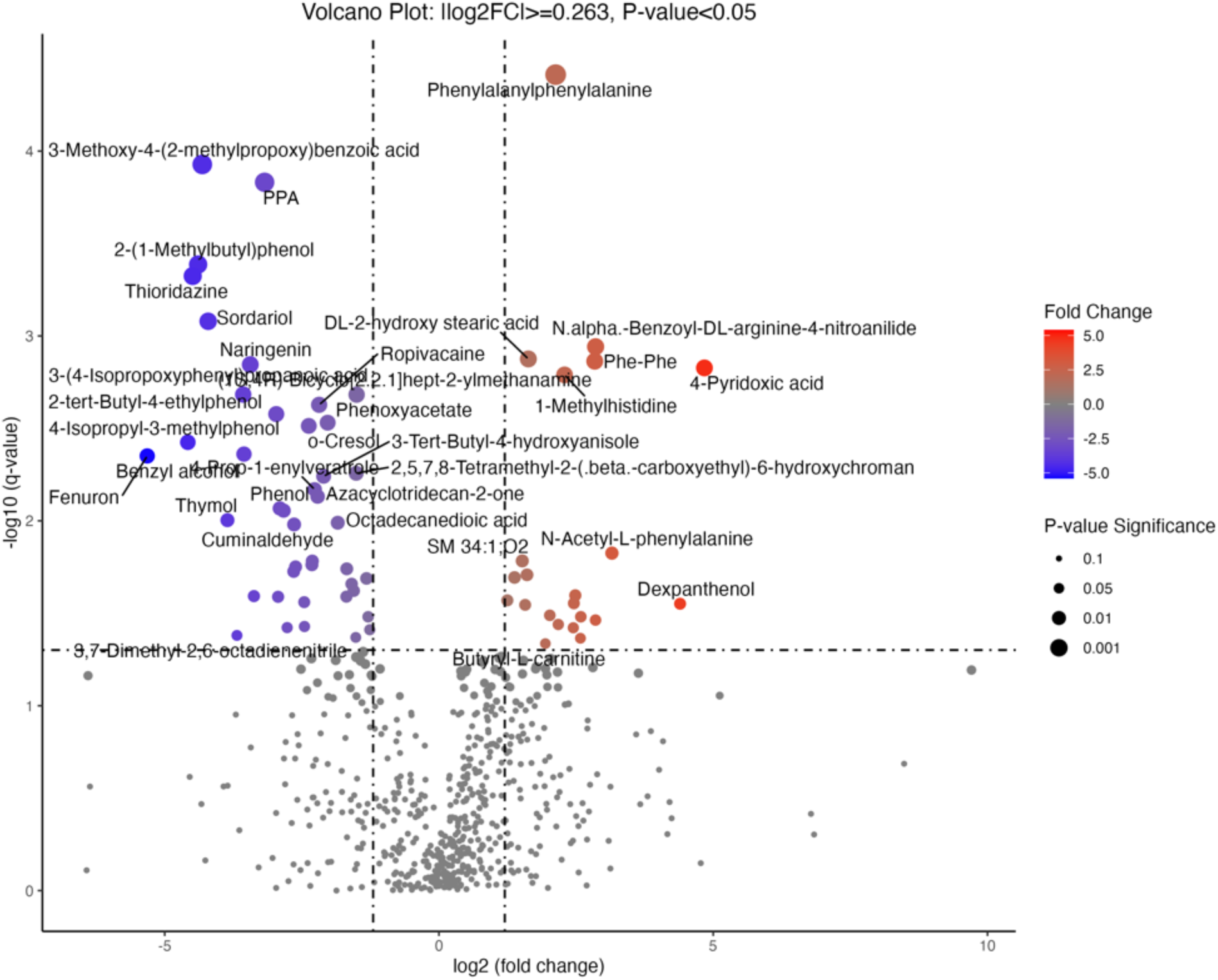
Volcano Plot Showing Statistically Significant Metabolite Changes Between OC II-III and OC IV Groups. Red dots represent metabolites upregulated in OC IV, while blue dots represent those upregulated in OC II-III. Fold change threshold: 1.2, raw p-value threshold: 0.05. Abbreviations: LPC, lysophosphatidylcholine; MG, Monoacylglycerol; Phe-Phe, phenylalanine-phenylalanine; Phe-Leu, phenylalanine-leucine; CerP, Ceramide phosphate; SM, sphingomyelin; LPI, Lysophosphatidylinositol; PC, Phosphatidylcholine; PPA, phenylpropionic acid; PE, Phosphatidylethanolamine.

**Figure S2.**
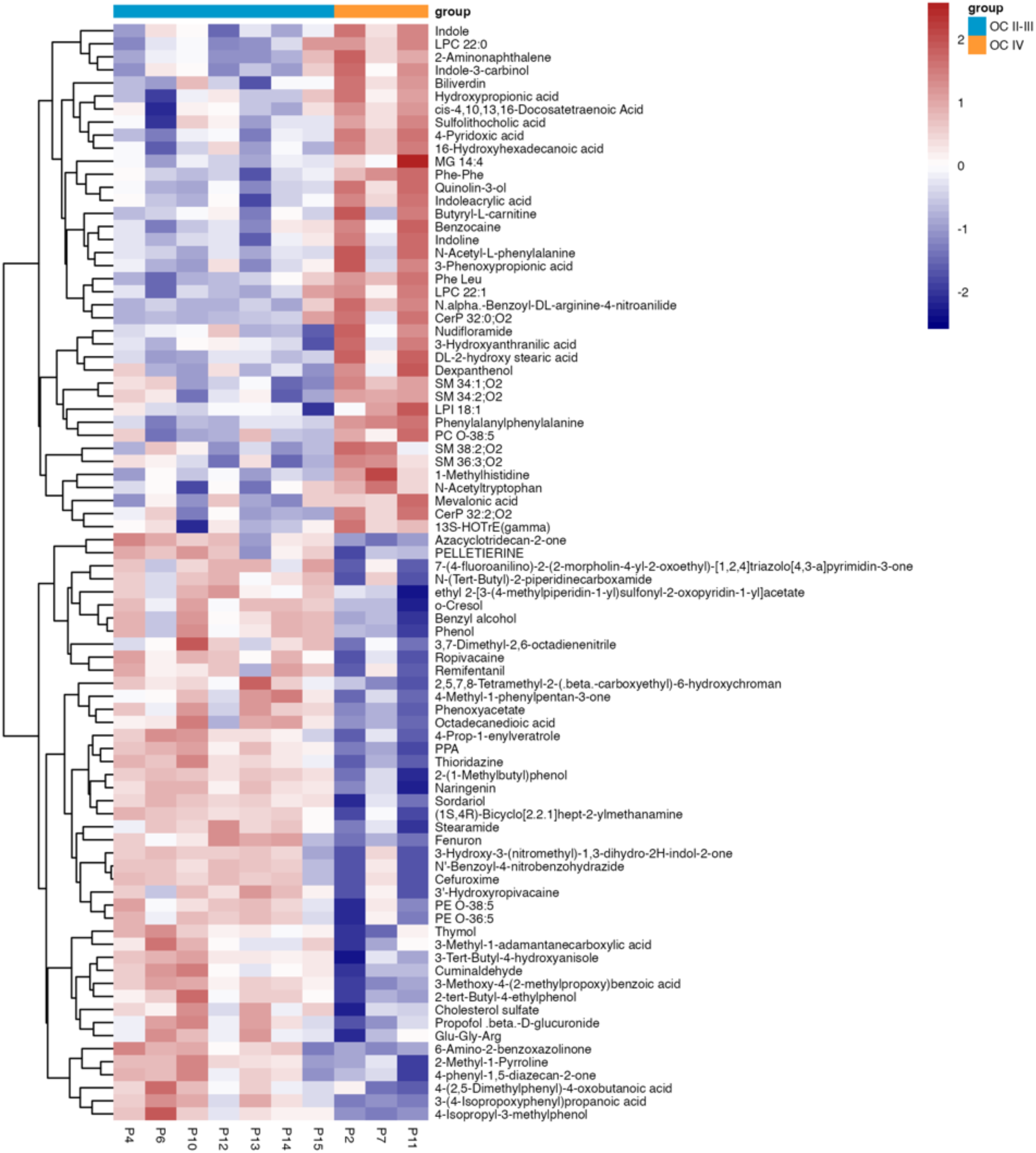
Identification of Ascites Metabolic Signatures in OC II-III and OC IV Groups. The heatmap displays 84 significant metabolites (t-test, raw p-value cutoff of 0.05, fold change threshold of 1.2) across seven OC II-III and three OC IV biological replicates. Clustering was performed using Ward’s hierarchical method with Euclidean distance as the distance metric. Abbreviations: LPC, lysophosphatidylcholine; MG, Monoacylglycerol; Phe-Phe, phenylalanine-phenylalanine; Phe-Leu, phenylalanine-leucine; CerP, Ceramide phosphate; SM, sphingomyelin; LPI, Lysophosphatidylinositol; PC, Phosphatidylcholine; PPA, phenylpropionic acid; PE, Phosphatidylethanolamine.

#### 3.5. Identification of microbiome-derived metabolite changes between OC stage II-III and OC stage IV

The same analysis using the MiMeDB database identified 16 metabolites potentially linked to the gut microbiome, including those associated with bacterial species, Eukaryota/Fungi, and Archaea. The corresponding phyla for the OC II-III vs. OC IV comparison are provided in **Table 3**.

**Table 3.**
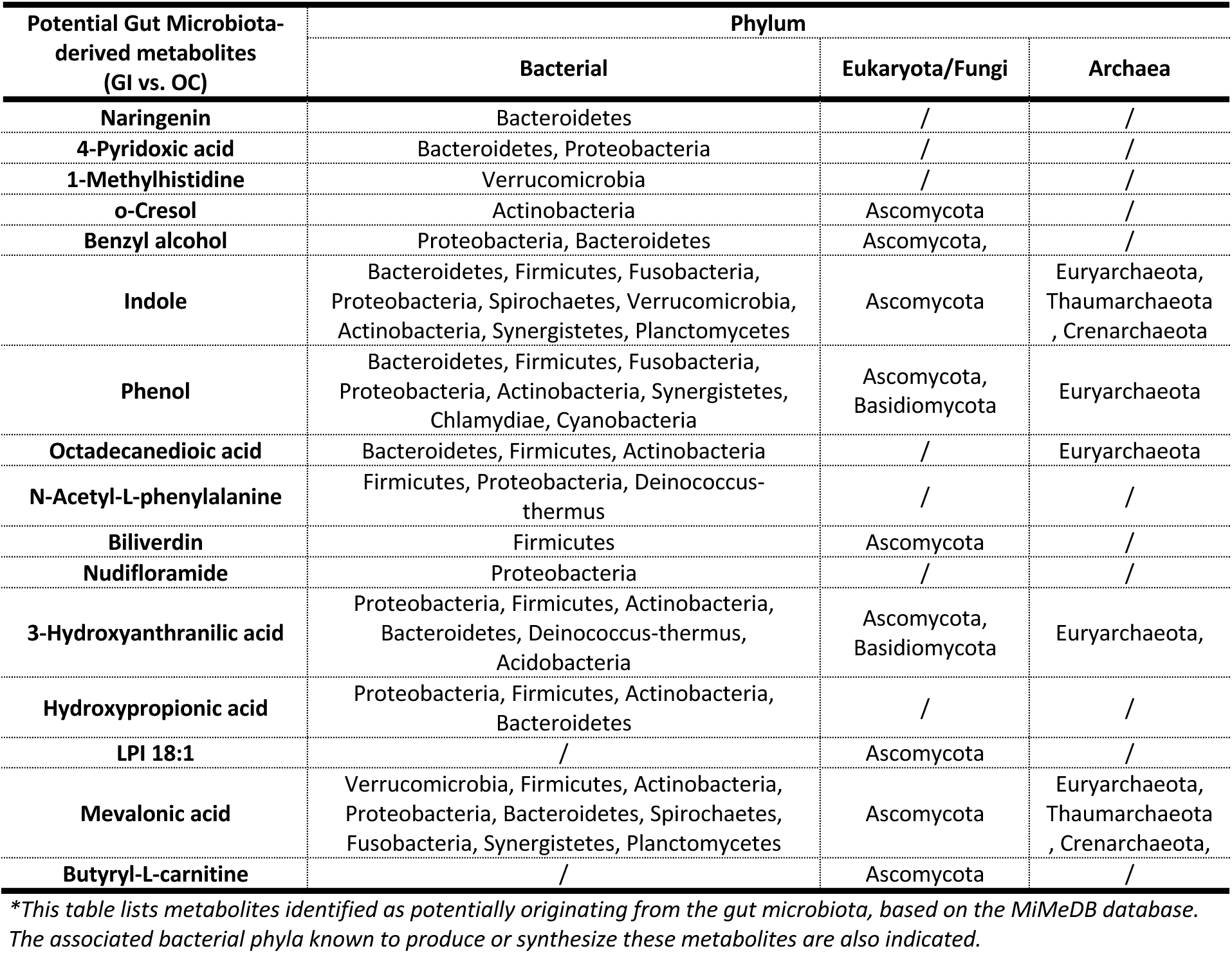
Metabolites Potentially Derived from the Gut Microbiota Identified in the Comparison Between Ovarian Cancer Stage II-III and Stage IV.

| Potential Gut Microbiota-derived metabolites (GI vs. OC) | Phylum |  |  |
| --- | --- | --- | --- |
|  | Bacterial | Eukaryota/Fungi | Archaea |
| Naringenin | Bacteroidetes | / | / |
| 4-Pyridoxic acid | Bacteroidetes, Proteobacteria | / | / |
| 1-Methylhistidine | Verrucomicrobia | / | / |
| o-Cresol | Actinobacteria | Ascomycota | / |
| Benzyl alcohol | Proteobacteria, Bacteroidetes | Ascomycota | / |
| Indole | Bacteroidetes, Firmicutes, Fusobacteria, Proteobacteria, Spirochaetes, Verrucomicrobia, Actinobacteria, Synergistetes, Planctomycetes | Ascomycota | Euryarchaeota, Thaumarchaeota, Crenarchaeota |
| Phenol | Bacteroidetes, Firmicutes, Fusobacteria, Proteobacteria, Actinobacteria, Synergistetes, Chlamydiae, Cyanobacteria | Ascomycota, Basidiomycota | Euryarchaeota |
| Octadecanedioic acid | Bacteroidetes, Firmicutes, Actinobacteria | / | Euryarchaeota |
| N-Acetyl-L-phenylalanine | Firmicutes, Proteobacteria, Deinococcus-thermus | / | / |
| Biliverdin | Firmicutes | Ascomycota | / |
| Nudifloramide | Proteobacteria | / | / |
| 3-Hydroxyanthranilic acid | Proteobacteria, Firmicutes, Actinobacteria, Bacteroidetes, Deinococcus-thermus, Acidobacteria | Ascomycota, Basidiomycota | Euryarchaeota, |
| Hydroxypropionic acid | Proteobacteria, Firmicutes, Actinobacteria, Bacteroidetes | / | / |
| LPI 18:1 | / | Ascomycota | / |
| Mevalonic acid | Verrucomicrobia, Firmicutes, Actinobacteria, Proteobacteria, Bacteroidetes, Spirochaetes, Fusobacteria, Synergistetes, Planctomycetes | Ascomycota | Euryarchaeota, Thaumarchaeota, Crenarchaeota, |
| Butyryl-L-carnitine | / | Ascomycota | / |

*\*This table lists metabolites identified as potentially originating from the gut microbiota, based on the MiMeDB database. The associated bacterial phyla known to produce or synthesize these metabolites are also indicated.*

The 16 differentiated potential gut microbiota-derived metabolites between OC II-III and OC IV showed significant changes. Specifically, 1-methylhistidine (A), 3-hydroxyanthranilic acid (B), 4-pyridoxic acid (C), biliverdin (E), butyryl-L-carnitine (F), hydroxypropionic acid (G), indole (G), lysophosphatidylinositol 18:1 (LPI 18:1) (I), mevalonic acid (J), N-acetyl-L-phenylalanine (K), and nudifloramide (M) were significantly elevated in OC IV ascites samples compared to OC II-III. On the other hand, benzyl alcohol (D), naringenin (L), o-cresol (N), octadecanedioic acid (O), and phenol (P) were significantly reduced in the OC IV group relative to OC II-III. These changes suggest distinct metabolic shifts between ovarian cancer stages II-III and IV, potentially reflecting the progression of the disease. The corresponding metabolite alterations are visualized in **Figure 7**.

**Figure 7.**
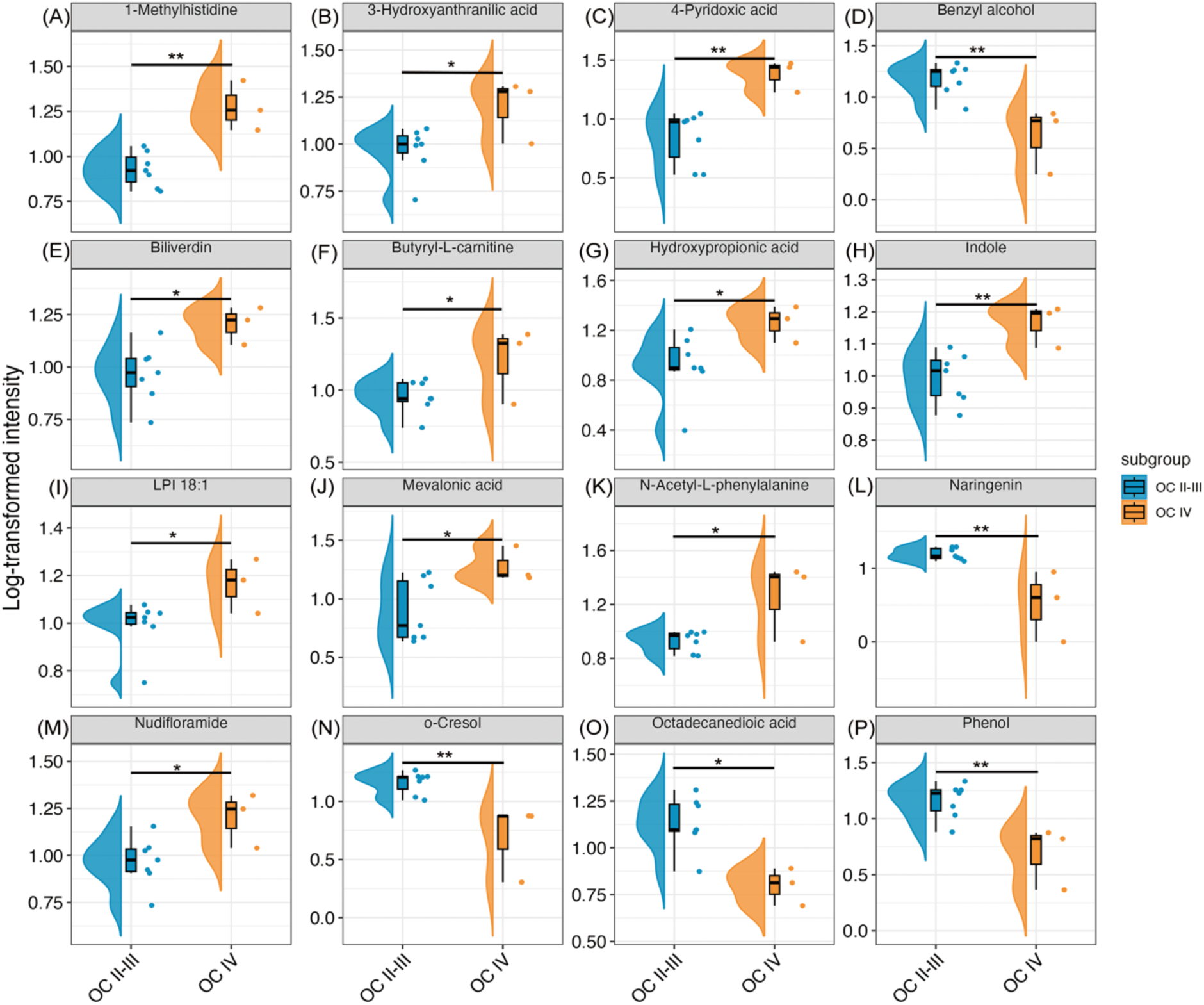
Raincloud Plots of Differentiated Potential Gut Microbiota-derived Metabolites Between OC II-III and OC IV. Raincloud plots display the significantly different metabolites: (A) 1-methylhistidine, (B) 3-hydroxyanthranilic acid, (C) 4-pyridoxic acid, (D) benzyl alcohol, (E) biliverdin, (F) butyryl-L-carnitine, (G) hydroxypropionic acid, (H) indole, (I) LPI 18:1 (lysophosphatidylinositol 18:1), (J) mevalonic acid, (K) N-acetyl-L-phenylalanine, (L) naringenin, (M) nudifloramide, (N) o-cresol, (O) octadecanedioic acid, and (P) phenol, comparing ovarian cancer stage II-III (OC II-III) with ovarian cancer stage IV (OC IV) groups. Unpaired t-tests were used for comparisons with normalized parametric data (*p < .05, **p < .01, ***p < .001, ****p < .0001). Dots represent individual data points, the middle line of the box represents the median, and the upper and lower edges of the box represent the upper and lower quartiles. Half-violins illustrate data distributions. Abbreviations: LPI, Lysophosphatidylinositol.

**Figure 8A** illustrates the broad metabolic variations in potential gut-derived metabolites across the 7 OC II-III and 3 OC IV ascites samples, highlighting distinct shifts between early and advanced stages of ovarian cancer. **Figure 8B** displays the microbial sources and categories of the metabolites that exhibited alterations between the OC II-III and OC IV groups, including their associated phyla and respective superkingdoms. Bacteria emerged as the primary source of these metabolites, followed by Eukaryota/Fungi, with Archaea contributing the least. These findings suggest that the gut microbiota’s involvement in ovarian cancer progression may stem from a diverse array of microbial species, each potentially playing a specific role in influencing the tumor microenvironment and disease progression.

**Figure 8.**
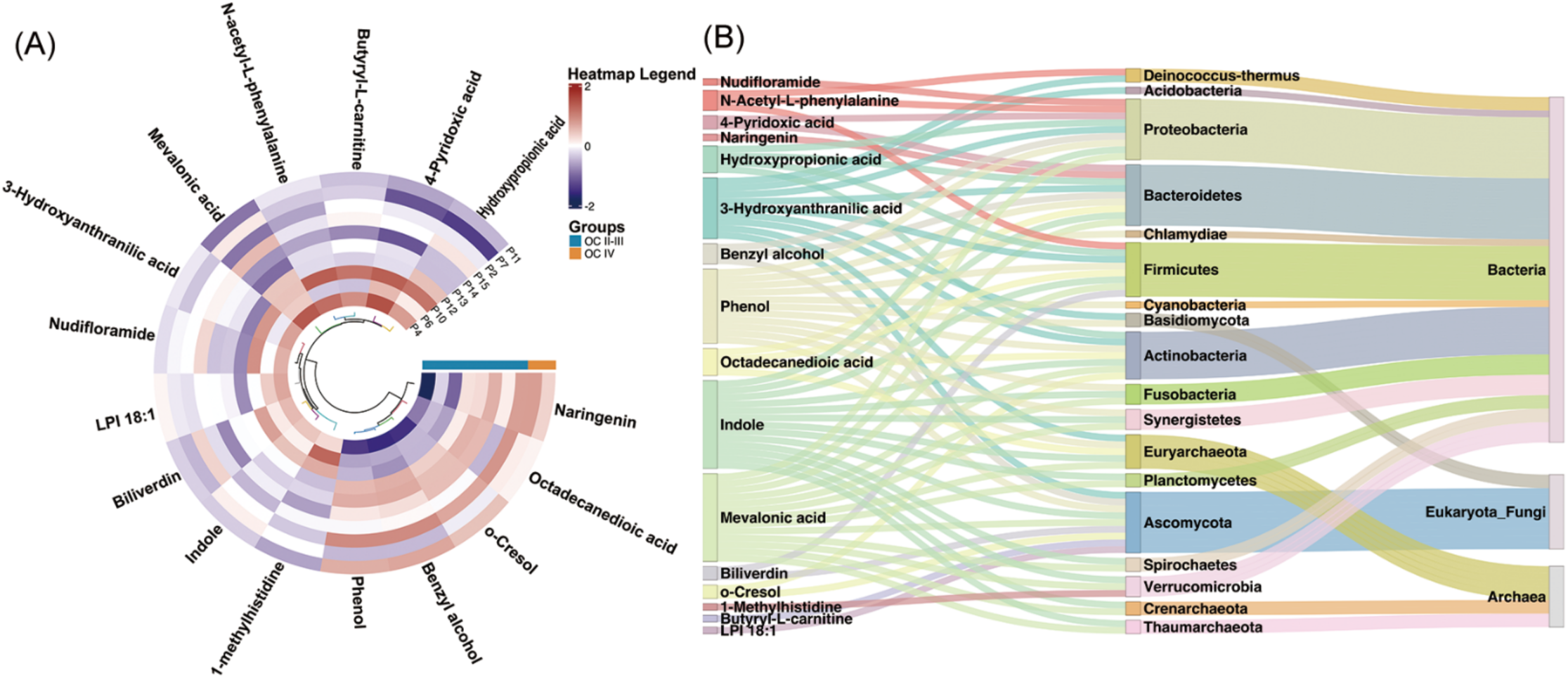
Intensity Variations of Potential Gut Microbiota-derived Metabolites Between OC II-III and OC IV Groups and Their Microbial Origins. (A) The circular heatmap illustrates the intensity variations of 16 significant gut microbiota-derived metabolites across 7 OC II-III and 3 OC IV biological replicates (t-test, raw p-value cutoff of 0.05, fold change threshold of 1.2). Clustering was performed using Ward’s hierarchical method with Euclidean distance. (B) The Sankey plot represents the microbial phyla and their respective superkingdoms linked to the altered metabolites, highlighting the dominant role of gut bacteria and their interactions with Eukaryota/Fungi and Archaea in the ascites of advanced ovarian cancer. Abbreviations: LPI, Lysophosphatidylinositol.

### 4. Correlations between potentially microbiota-derived metabolites and cytokines in Ascites

#### 4.1. Ovarian Cancer VS. Gastrointestinal Cancer

To investigate potential interactions between gut microbiota-derived metabolites and cytokines, a correlation analysis was performed between individual metabolites from the OC vs. GI comparison and cytokine levels in **Figure 9**. The results revealed several significant correlations. IL-23 showed a positive correlation with D-glucurono-6,3-lactone and a negative correlation with trimethylamine N-oxide. IL-18 was negatively correlated with caffeine. IL-10 demonstrated significant positive correlations with glucosamine, D-tagatose, trimethylamine N-oxide, caffeine, LPC 22:4, and LPC 20:1, while showing negative correlations with benzamide and thymol. MCP-1 was negatively correlated with D-tagatose. TNF-α positively correlated with D-glucurono-6,3-lactone. IFN-γ was negatively correlated with levulinic acid and trimethylamine N-oxide, while IFN-α2 showed a positive correlation with D-glucurono-6,3-lactone.

**Figure 9.**
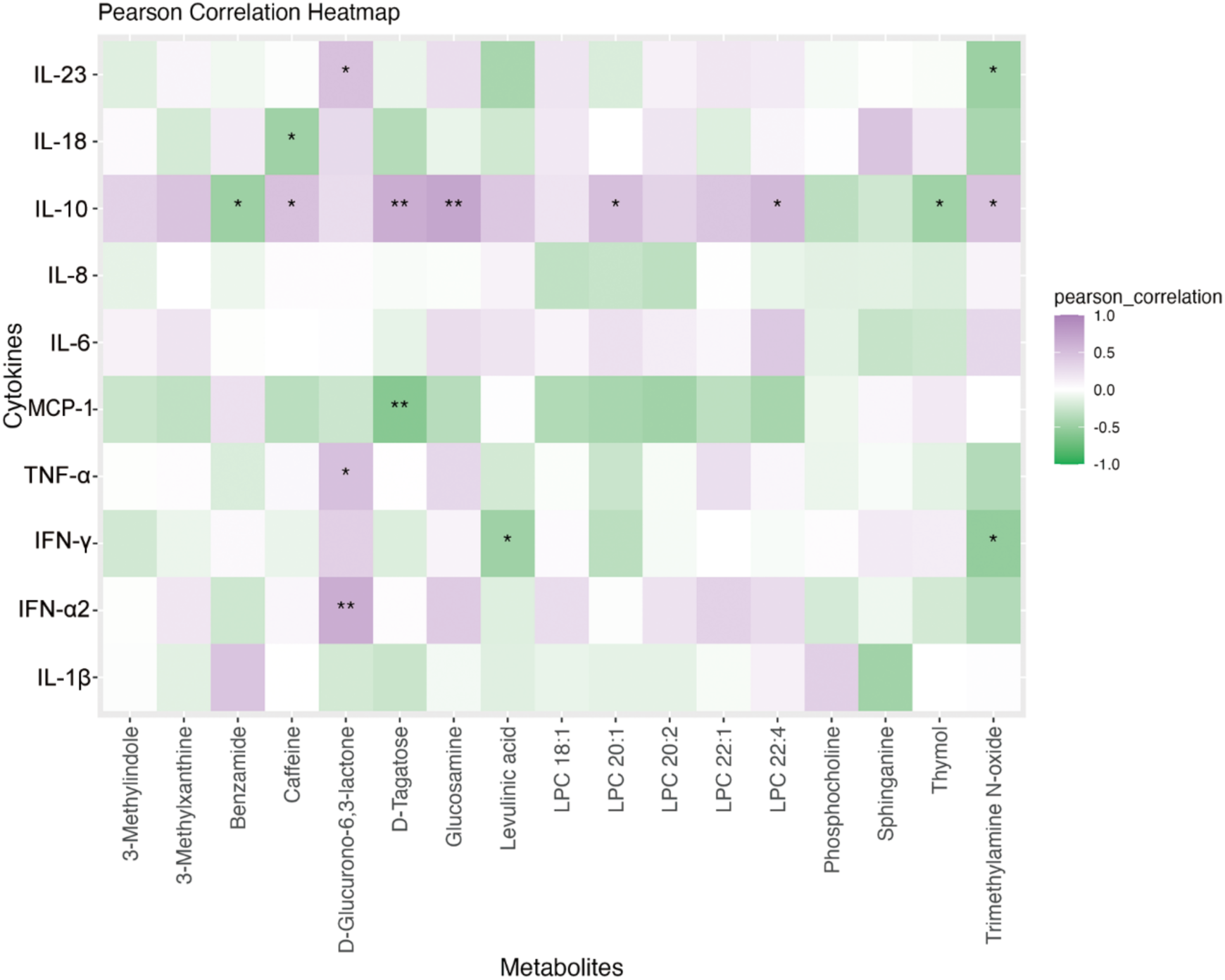
Heatmap of Correlations Between Potential Gut Microbiota-derived Metabolites (OC vs. GI) and Flow Cytometry Cytokines. The heatmap illustrates the correlations between gut microbiota-derived metabolites and cytokines measured by flow cytometry. A single asterisk (*) indicates a significant correlation (p < 0.05), while a double asterisk (**) represents a highly significant correlation (p < 0.01). Correlations were calculated using a combination of Pearson and Spearman methods. Abbreviations: LPC, lysophosphatidylcholine.

#### 4.2. Ovarian Cancer Stage II-III VS. Stages IV

The same correlation analysis was conducted between individual gut microbiota-derived metabolites from the OC II-III vs. OC IV comparison and cytokine levels (**Figure 10**). The analysis revealed significant negative correlations between IL-8 and both 3-hydroxyanthranilic acid and nudifloramide. IL-6 showed significant negative correlations with butyryl-L-carnitine, indole, and N-acetyl-L-phenylalanine. MCP-1 was positively correlated with benzyl alcohol, naringenin, o-cresol, octadecanedioic acid, and phenol, while showing significant negative correlations with 1-methylhistidine, 4-pyridoxic acid, and mevalonic acid. IL-1B displayed positive correlations with 1-methylhistidine, 4-pyridoxic acid, LPI 18:1, mevalonic acid, N-acetyl-L-phenylalanine, and nudifloramide, while significantly negatively correlated with benzyl alcohol, naringenin, o-cresol, octadecanedioic acid, and phenol.

**Figure 10.**
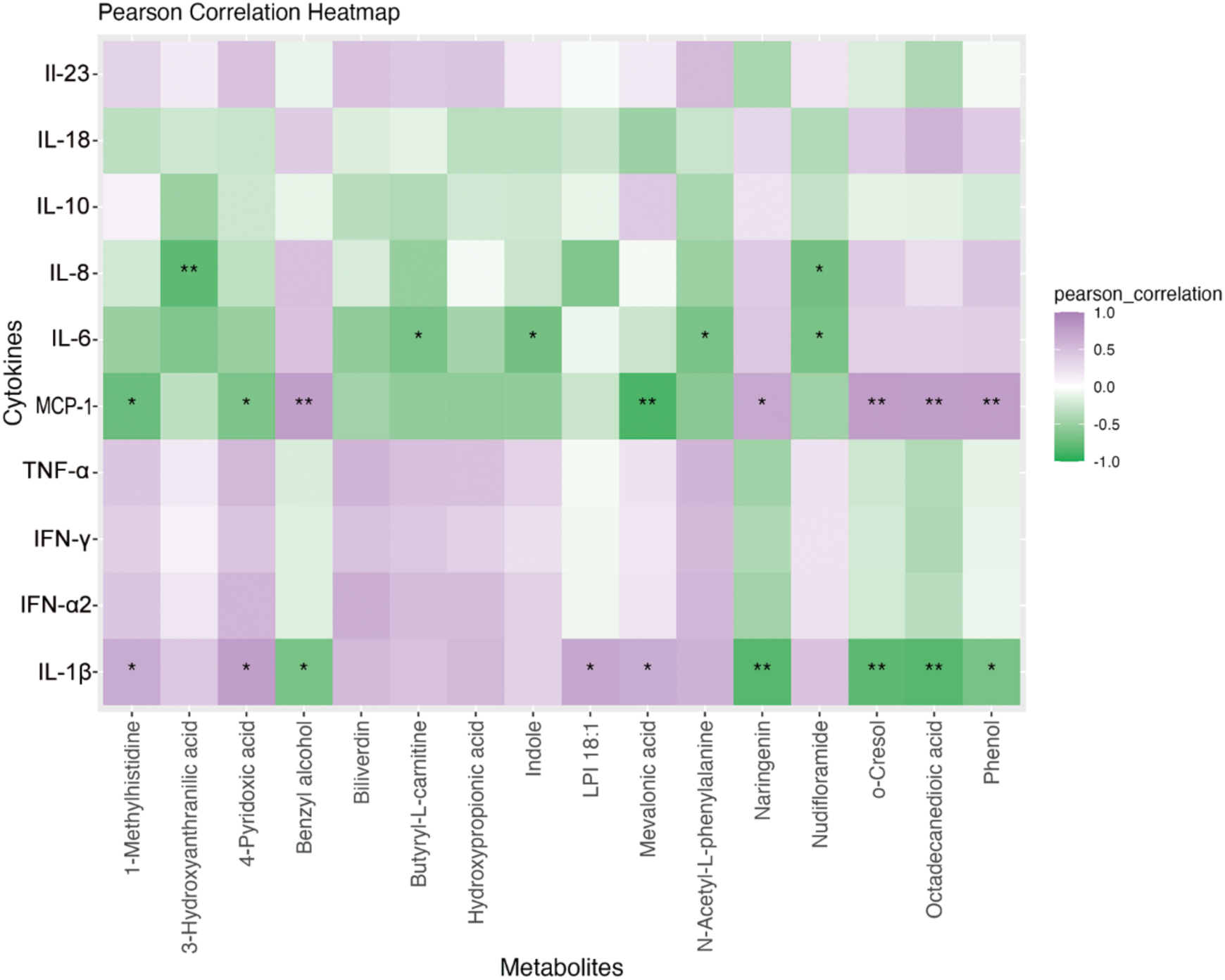
Heatmap of Correlations Between Potential Gut Microbiota-derived Metabolites (OC II-III vs. OC IV) and Flow Cytometry Cytokines. The heatmap illustrates the correlations between gut microbiota-derived metabolites and cytokines measured by flow cytometry. A single asterisk (*) indicates a significant correlation (p < 0.05), while a double asterisk (**) represents a highly significant correlation (p < 0.01). Correlations were calculated using a combination of Pearson and Spearman methods. Abbreviations: LPI, Lysophosphatidylinositol.

## Discussion

Malignant ascites forms due to a combination of increased fluid production and reduced lymphatic absorption(24, 25). Normally, the peritoneum absorbs excess fluid through lymphatic channels, but in malignancy, the tumor’s growth and invasion disrupt these processes. Tumor-induced neovascularization(7), driven by vascular endothelial growth factor (VEGF), increases vascular permeability, leading to fluid leakage into the abdominal cavity(8, 26). Matrix metalloproteinases (MMPs) also degrade tissue barriers, further promoting fluid accumulation(27, 28). In addition, hormonal changes activate the renin-angiotensin-aldosterone system, causing sodium and fluid retention(6).

Emerging research suggests the gut microbiota may influence ascites development. Disruption of gut bacteria has been linked to inflammation, altered immune responses, and cancer progression(29-31). In malignancies, an imbalanced gut microbiota can impair the intestinal barrier, increasing permeability and allowing bacterial translocation(32, 33). This worsens inflammation and fluid retention, creating a cycle that perpetuates ascites formation. The role of gut microbiota in modulating immune and metabolic pathways presents a potential area for further exploration in managing malignant ascites(34).

### LC-MS based metabolomic profiles of OC stages II-III, IV and GI ascites samples

Metabolic analysis of ascites from the OC II-III, OC IV, and GI groups revealed that OC IV shares more similarities with the GI group, while OC II-III is distinctly different from both. This pattern aligns with the progression of ovarian cancer, where advanced stages involve cancer cells detaching from the primary tumor, surviving in the peritoneal fluid, and spreading to organs such as the liver, lungs, spleen, intestines, and lymph nodes(35-37).

One metabolite, phenylalanylphenylalanine, a dipeptide of two phenylalanine molecules, was found at higher levels in OC IV. Elevated phenylalanine levels and altered phenylalanine-to-tyrosine ratios have been associated with inflammatory conditions, including cancer(38). Phenylalanine metabolism also influences T-cell function, regulate T-cell proliferatio and activation and affecting the following immune response(38). This suggests that higher levels of phenylalanylphenylalanine could be linked to more advanced cancer stages. Similarly, SM 36:3-O2, a sphingomyelin, was significantly elevated in OC IV. Increased sphingomyelin levels have been linked to cancer development, with altered sphingomyelin metabolism observed in metastatic tumor cells and various cancers(39-41). These changes in sphingomyelin metabolism, including higher synthesis and reduced breakdown, contribute to disrupted lipid balance, promoting cancer growth, particularly in ovarian and breast cancers(42, 43). The higher levels of sphingomyelin in ascites may, therefore, reflect more aggressive tumor behavior and advanced disease.

LC-MS analysis also detected a wide range of metabolites in the ascites samples, many of which are not directly related to human metabolism. Several of the significantly different compounds were identified as exogenous substances, including drugs, food-derived compounds, plant oil-derived, and anesthetics. These findings highlight the complexity of ascites as a mixture of both endogenous and exogenous compounds. The presence of these exogenous compounds can be explained by several factors. Medications such as chemotherapeutics, anesthetics, and pain management drugs are commonly used in ovarian cancer treatment, and residual traces can accumulate in bodily fluids(44). Additionally, diet and environmental exposure significantly influence an individual’s metabolic profile(45, 46).

Plant-derived compounds or food additives can enter the bloodstream and appear in ascites, particularly in patients undergoing systemic changes due to disease or treatment(47). This suggests that ascites is influenced by both internal metabolic processes and external factors, complicating the interpretation of LC-MS data.

### Potential microbiota-derived metabolomic features in malignant ascites samples

Through MiMeDB analysis, we identified 17 gut microbiota-derived metabolites in the OC vs. GI comparison and 16 in the OC II-III vs. OC IV comparison. Both sets revealed a predominance of bacterial-origin metabolites in malignant ascites, consistent with the human gut microbiota profile(48). While the proportions of Archaea and Eukaryota/Fungi-derived metabolites showed slight differences between the two sets, the findings align with studies highlighting the role of gut microbiota in carcinogenesis, immune surveillance, and responses to immunotherapy(49-51).

In OC patients, continuous Immune checkpoint blockade (ICB) therapy with poly (ADP-ribose) polymerase inhibitors (PARPi) has demonstrated efficacy in prolonging progression-free and overall survival(52-55). For GI cancers, ICB strategies vary by tumor origin. Immunotherapy is now standard in first-line treatment for advanced colorectal cancer with high microsatellite instability(56-59). Additionally, in advanced gastric cancer, combining immunotherapy with chemotherapy or with HER2-targeted therapy has shown significant and lasting survival benefits in HER2-positive patients(60-63).

Despite advances in immunotherapy, a significant proportion of patients exhibit primary or acquired resistance to treatment(64, 65). Additionally, immunotherapy-related adverse reactions pose a clinical challenge, particularly with the expanded use of combination therapies and multi-agent immunotherapy(66-73). To address these challenges, studies have identified host-associated genomic and molecular biomarkers predictive of immunotherapy response(74-76). Emerging evidence also implicates the gut microbiome, particularly specific microbial taxa, in modulating immune checkpoint blockade (ICB) efficacy(77).

Research suggests that gut microbiome composition may be both predictive and prognostic of therapeutic response to ICB, highlighting its potential as a biomarker(77). These insights have driven the development of microbiome-targeted strategies aimed at enhancing treatment efficacy and minimizing adverse effects by modulating the patient ’ s gut microbiota(78, 79).

The gut microbiota-derived metabolites identified in ascites contribute significantly to the tumor microenvironment. This study revealed distinct microbiota profiles across ovarian cancer stages and gastrointestinal cancers, highlighting the importance of larger sample sizes and advanced tools like 16S rRNA sequencing to enhance our understanding. Further research is needed to explore how these microbial profiles correlate with immunotherapy side effects, tumor reduction efficacy, and clinical outcomes, providing insight into their role in ascites formation and cancer progression.

Emerging interventions, such as fecal microbiota transplants (FMT), prebiotics, probiotics, antibiotics, and dietary modifications, show promise in modulating the gut microbiome(80-82). Characterizing gut microbiota and its systemic effects will be key to identifying actionable targets for future therapeutic interventions and clinical assessment.

### Microbiota-Derived metabolomic profiles in ascites from OC and GI

Several bacterial-derived metabolites identified in the OC vs. GI comparison are associated with immune-metabolic pathways and may affect immune responses in ovarian and gastrointestinal cancers. Lysophosphatidylcholines (LPCs), known pro-inflammatory lipids(83), are influenced by microbial taxa such as Bacteroidetes and firmicutes(84). Bacteroidetes contribute to lipid absorption and metabolism, while firmicutes play a role in immune modulation through lipid pathways(85-87). In cancer, LPCs contribute to inflammation, tumor growth, and immune evasion(88). Dysbiosis between these taxa may alter LPC levels and functions, impacting cancer progression, immune response, and gut health. The presence of LPCs (LPC 18:1, LPC 20:1, LPC 20:2, LPC 22:1, LPC 22:4) and sphingolipids (sphinganine) indicates significant involvement in membrane turnover and lipid signaling, both commonly disrupted in cancer(89-91).

Additionally, metabolites such as 3-methylindole and trimethylamine N-oxide suggest shifts in gut microbiota metabolism that may impact cancer progression or immune function(92, 93). Detoxification metabolites, like D-glucurono-6,3-lactone, reflect an active response to cellular stress, arising from cancer or external treatments such as chemotherapy(94).

### Differential potential microbiota-derived Metabolites in ascites: OC II-III vs. OC IV

In the comparison between OC stages II-III and IV, several metabolites reflect metabolic changes, immune modulation, and inflammatory responses typical of the tumor microenvironment, particularly in advanced stages. Metabolites such as mevalonic acid, butyryl-L-carnitine, and LPI 18:1 suggest shifts in lipid metabolism, likely driven by heightened energy demands, membrane synthesis, and signaling activity in ovarian cancer cells(95, 96). Additionally, 3-hydroxyanthranilic acid, indole, and naringenin indicate the presence of immune-modulating and inflammatory metabolites, potentially promoting immune evasion and supporting a pro-inflammatory environment in advanced cancer(97-100).

LPI 18:1, a bioactive lipid involved in cell signaling, is linked to tumor growth, migration, and immune suppression, and its levels may be influenced by gut microbiota, especially in cases of dysbiosis(95, 96). Indole, a microbial byproduct of tryptophan degradation, can affect immune responses and inflammation, potentially aiding immune evasion in cancer(99). These findings underscore the interaction between gut microbial metabolites and the tumor microenvironment, highlighting the gut-tumor axis’s role in cancer progression and suggesting therapeutic strategies that target the microbiota to improve outcomes.

Naringenin, known for its anti-inflammatory, antioxidant, and anticancer effects(100-102), was elevated in OC II-III compared to OC IV. It modulates inflammation by suppressing cytokine production and enhancing cytokine degradation(103), while also regulating cell growth, apoptosis, and metastasis(104, 105). Its higher levels in OC II-III suggest naringenin may help regulate immune responses and inhibit cancer progression in early stages, with reduced activity as the disease advances.

### Interactions between microbiota-derived metabolites and cytokines/chemokines

To investigate how gut microbiota-derived metabolites interact with the immune landscape in cancer progression, we conducted correlation analyses between these metabolites and cytokines in OC vs. GI and OC stage II-III vs. IV comparisons. The anti-inflammatory cytokine IL-10 (**Figure 9**) showed multiple positive correlations with metabolites such as glucosamine, D-tagatose, TMAO, caffeine, LPC 22:4, and LPC 20:1, suggesting these metabolites may contribute to immune suppression in the tumor microenvironment. This immune tolerance could facilitate tumor evasion from immune surveillance. Conversely, IL-10 was negatively correlated with benzamide and thymol, illustrating how different metabolites exert opposing influences on cytokine regulation. Notably, IL-10 in ascites has been associated with both the migration of ovarian cancer cells(106)and, in some cases, longer survival in patients receiving cell-free and concentrated ascites reinfusion therapy (CART)(107).

MCP-1 (**Figure 10**), a chemokine that recruits monocytes and macrophages to the tumor site, was positively correlated with metabolites like benzyl alcohol, naringenin, o-cresol, octadecanedioic acid, and phenol, potentially promoting immune cell recruitment and inflammation. In contrast, 1-methylhistidine, 4-pyridoxic acid, and mevalonic acid were negatively correlated with MCP-1, potentially reducing immune cell recruitment. This dual influence suggests a complex balance of pro- and anti-inflammatory signals in advanced ovarian cancer, where shifts in metabolite levels may influence MCP-1 activity. Additionally, MCP-1’s positive association with infertility in endometriosis patients highlights its role in inflammatory immune reactions within the peritoneal cavity(108-110).

Together, these findings illustrate an intricate network of interactions between metabolites and cytokines that likely impact cancer progression, immune evasion, and patient outcomes. Observed stage-dependent differences underscore the influence of metabolic shifts on immune responses within the tumor microenvironment. The balance of pro- and anti-inflammatory metabolites, in particular, may play a critical role in shaping this environment.

For future studies, expanding the sample size would improve statistical robustness and support more definitive conclusions on metabolite variations across cancer stages. The detection of bacterial metabolites could also be enhanced by integrating LC-MS with targeted platforms like 16S rRNA sequencing, which would refine the characterization of microbial contributions to the metabolome. This combined approach would deepen understanding of the microbiome’s role in ascites and its implications for cancer progression.

To visually summarize our findings, especially the gut microbiota-derived metabolomic differentiation in ascites samples from OC II-III and OC IV, we created a graphical abstract(**Figure 11**).

**Figure 11.**
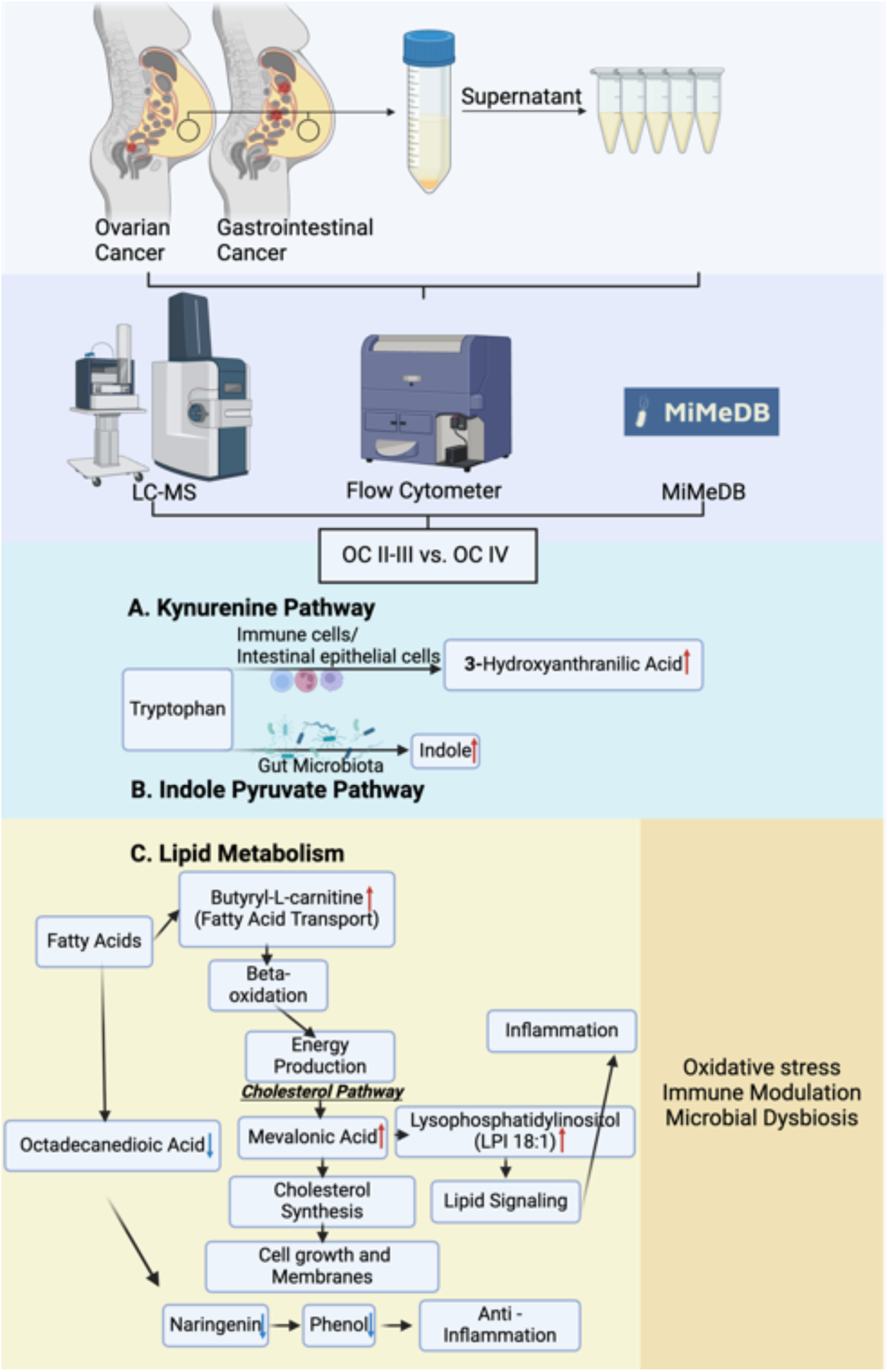
Graphical abstract to present gut microbiota-derived metabolomic differentiation in ascites samples from OC II-III and OC IV. Red and blue arrow means increase and decrease in OC IV ascites compare to OC II-III.

## Conclusion

This study utilized novel trapped ion mobility time of flight (timsTOF) mass spectrometry to profile metabolites in malignant ascites samples, comparing gastrointestinal cancers and different stages ovarian cancers. Both reversed-phase and hydrophilic interaction liquid chromatography were employed to ensure comprehensive separation of non-polar and polar metabolites. Key findings revealed distinct potential microbiota-derived metabolic changes in OC versus GI cancers and across ovarian cancer stages, particularly involving lipid metabolism, with significant alterations in sphingolipid and phospholipid pathways. Additionally, potential gut microbiota-derived metabolites were identified and correlated with cytokine and chemokine levels, indicating a possible interaction between gut microbiota and the immune response in ovarian and gastrointestinal cancers. These findings show the presence of microbiota-derived metabolites in malignant ascites and contribute as first step to a better understanding of the interplay between microbiota and intestinal malignancies. Future studies upon malignant ascites shall follow, assessing the exact impact of individual microbial metabolites upon the tumor micro environment in order to improve patient care and support the success of novel cancer therapies.

## Contributions

Conception: C.T., S.D., N.B., S.K., A.M.

Data acquisition: W.K., Y.S., G.B., Q.Y., K.C.

Data analysis: W.K., S.D., C.T.

Interpretation of data: W.K., C.T., S.D.

Figure preparation: S.D.

Manuscript draft: S.D.

Manuscript editing: S.K., N.B., J.S., A.M., C.T.

All authors have approved the submitted version.

S.D., W.K., K.C, Q.Y., Y.S., G.B., S.K., N.B., J.S., A.M., C.T.

## Data availability statement

The data supporting this study are available from the corresponding author upon reasonable request.

## Abbreviations

9-OAHSA: 9-Oxoheptadecanoic Acid
ADENOCA: Subtype unknown adenocarcinoma
Ala Glu Ile Lys: Alanine-Glutamic Acid-Isoleucine-Lysine peptide
CCS: Collision cross section
Cer 42:2;O2: Ceramide with 42 carbons, 2 double bonds, and 2 oxygen atoms
CerP 28:1;O2: Ceramide Phosphate with 28 carbons, 1 double bond, and 2 oxygen atoms
CerP 32:0;O2: Ceramide Phosphate with 32 carbons and no double bonds
Co: Control
DG 34:3: Diacylglycerol with 34 carbons and 3 double bonds
GC: Gas chromatography
Glu-Gly-Arg: Glutamic Acid-Glycine-Arginine peptide
HGSC: High-grade serous carcinoma
HILIC: Hydrophilic interaction liquid chromatography
HMDB: Human Metabolome Database
HPLC: High-performance liquid chromatography
IL-10: Interleukin 10
IL-18: Interleukin 18
IL-6: Interleukin 6
IL-8: Interleukin 8
LC: Liquid chromatography
Leu Leu Val Val Ala: Leucine-Leucine-Valine-Valine-Alanine peptide
LPC: Lysophosphatidylcholine
LPC 22:0: Lysophosphatidylcholine with a 22-carbon saturated fatty acid
LPC 22:1: Lysophosphatidylcholine with a 22-carbon monounsaturated fatty acid
LPC 22:2: Lysophosphatidylcholine with a 22-carbon polyunsaturated fatty acid
LPC 22:3: Lysophosphatidylcholine with a 22-carbon fatty acid and 3 double bonds
LPE: Lysophosphatidylethanolamine
LPE O-16:0: Lysophosphatidylethanolamine with a 16-carbon saturated fatty acid and an ether linkage
LPE O-16:1: Lysophosphatidylethanolamine with a 16-carbon fatty acid and 1 double bond, with an ether linkage
LPE O-16:2: Lysophosphatidylethanolamine with a 16-carbon fatty acid and 2 double bonds, with an ether linkage
LPE O-18:2: Lysophosphatidylethanolamine with an 18-carbon fatty acid and 2 double bonds, with an ether linkage
LPE O-18:3: Lysophosphatidylethanolamine with an 18-carbon fatty acid and 3 double bonds, with an ether linkage
LPS 18:1: Lysophosphatidylserine with an 18-carbon fatty acid and 1 double bond
LPS 20:0: Lysophosphatidylserine with a 20-carbon saturated fatty acid MCP-1 Monocyte chemoattractant protein-1
MG 18:2: Monoglyceride with an 18-carbon fatty acid and 2 double bonds MQ MilliQ water
MS: Mass spectrometry
NAFLD: Non-alcoholic fatty liver disease
NIST: National Institute of Standards and Technology
NMR: Nuclear magnetic resonance
OC III: Ovarian cancer stage III
OC IV: Ovarian cancer stage IV
PASEF: Parallel Accumulation–Serial Fragmentation
PC: Phosphatidylcholine
PC O-38:5: Phosphatidylcholine with 38 carbons, 5 double bonds, and an ether linkage
PE: Phosphatidylethanolamine
PE 38:1: Phosphatidylethanolamine with 38 carbons and 1 double bond
PE O-34:3: Phosphatidylethanolamine with 34 carbons, 3 double bonds, and an ether linkage
PE O-36:5: Phosphatidylethanolamine with 36 carbons, 5 double bonds, and an ether linkage
PE O-38:5: Phosphatidylethanolamine with 38 carbons, 5 double bonds, and an ether linkage
PE O-38:6: Phosphatidylethanolamine with 38 carbons, 6 double bonds, and an ether linkage
PG 18:1_18:2: Phosphatidylglycerol with one 18:1 and one 18:2 fatty acid chains Phe Leu Phenylalanine-Leucine peptide
Phe-Phe: Phenylalanine-Phenylalanine dipeptide
PPA: Phenylpropionic Acid
PQN: Probabilistic quotient normalization
QTOF: Quadrupole Time-of-Flight
RNS: Nitrogen species
ROS: Reactive oxygen species
RPLC: Reversed-phase liquid chromatography
SBP: Spontaneous bacterial peritonitis
SM 30:1;O2: Sphingomyelin with 30 carbons, 1 double bond, and 2 oxygen atoms SM 30:2;O2 Sphingomyelin with 30 carbons, 2 double bonds, and 2 oxygen atoms
SM 32:1;O2: Sphingomyelin with 32 carbons, 1 double bond, and 2 oxygen atoms SM 32:2;O2 Sphingomyelin with 32 carbons, 2 double bonds, and 2 oxygen atoms
SM 34:1;O2: Sphingomyelin with 34 carbons, 1 double bond, and 2 oxygen atoms
SM 34:1;O2: Sphingomyelin with 34 carbons, 1 double bond, and 2 oxygen atoms
SM 34:2;O2: Sphingomyelin with 34 carbons, 2 double bonds, and 2 oxygen atoms
SM 36:3;O2: Sphingomyelin with 36 carbons, 3 double bonds, and 2 oxygen atoms
SM 44:6;O2: Sphingomyelin with 44 carbons, 6 double bonds, and 2 oxygen atoms
TAM: Tumor-associated macrophage
TIMS: Trapped Ion Mobility Spectrometry
VIP-HESI: Vacuum insulated probe heated electrospray ionization

